# Development of a prediction model for infant hospitalization and death using clinical features assessed by community health workers during routine postnatal home visits in Dhaka, Bangladesh

**DOI:** 10.1101/2025.10.19.25338306

**Authors:** Alastair Fung, Marimuthu Sappani, Cole Heasley, Chun-Yuan Chen, Shaun K. Morris, Peter J. Gill, Diego G. Bassani, Davidson H. Hamer, Prakesh S. Shah, S. M. Abdul Gaffar, Sultana Yeasmin, Shafiqul A. Sarker, Shamima Sultana, Joseph Beyene, Daniel E. Roth

## Abstract

**Introduction:** To improve upon the World Health Organization (WHO) 8 danger signs used to identify young infants (<2 months) requiring referral during community health worker (CHW) home visits, aggregative features (e.g., cumulative visits with fever) rather than visit-specific features (e.g., fever at a single visit), and a machine learning random forest model, may enhance predictive performance. Applying these approaches, we aimed to develop a prediction model for infant hospitalization and/or death using CHW-assessed clinical features during home visits in Dhaka, Bangladesh.

**Methods:** We analyzed data from generally healthy infants prospectively enrolled at birth and assessed at 11 scheduled CHW visits from 3-60 days of age. To predict first hospitalization or death, we developed two models – time-varying Cox regression and random forest – using the same set of candidate predictors (45 clinical features of which 8 were WHO danger signs, and 12 additional covariates) with aggregative features incorporated. We evaluated discrimination (C-statistic) and calibration (calibration plots). Performance was compared to a time-varying Cox model using only WHO danger signs.

**Results:** Among 1906 infants, 176 (9.2%) had an event (173 hospitalizations, 3 deaths). The best-performing Cox model (C-statistic=0.71; 95% CI 0.68-0.75) consisting of three baseline covariates (any perinatal/delivery complication, umbilical cord care, gestational age) and four visit-specific clinical features (nasal congestion, cough, jaundice, skin rash), and a Cox model with these four features plus WHO danger signs (C-statistic=0.70; 95% CI 0.67-0.74), demonstrated higher discrimination than WHO danger signs alone (C-statistic=0.56; 95% CI 0.54-0.60), with similar calibration. A random forest model (42 predictors) was well-calibrated with comparable discrimination (C-statistic=0.69; 95% CI 0.64-0.73).

**Conclusion:** Aggregative features and random forest did not outperform a time-varying Cox model using baseline covariates and visit-specific features. Adding four features to WHO danger signs may improve predictive performance by capturing a broader spectrum of infant illnesses requiring hospitalization.

**What is already known on this topic:**

- During community health worker (CHW) home visit assessments of young infants (<2 months), use of World Health Organization (WHO)-recommended danger signs to predict hospitalization and/or death may have limited sensitivity and may miss cases of severe illness requiring referral.
- Summarizing repeated assessments of clinical features during sequential home visits as aggregative predictors (e.g., cumulative visits with fever) rather than visit-specific predictors (e.g., fever at a single visit), and machine learning models such as random forest, have not been previously evaluated for prediction of infant hospitalization and/or death and may improve predictive performance compared to WHO danger signs.

**What this study adds:**

- Random forest, and use of aggregative predictors in both a random forest model and a time-varying Cox model, did not improve prediction of infant hospitalization and/or death during CHW routine home visits compared to a time-varying Cox model consisting of baseline covariates and visit-specific clinical features.
- Adding four visit-specific clinical features to the WHO danger signs improved prediction of hospitalization and/or death during CHW routine home visit assessments of young infants born generally healthy in an urban setting.

**How this study might affect research, practice or policy:**

- The findings support future research evaluating whether adding visit-specific clinical features to the WHO danger signs algorithm can improve identification of infants needing referral across diverse settings and with varying baseline risks.

## INTRODUCTION

In 2023, 2.3 million infants worldwide died in the first month of age, with the highest burden in sub-Saharan Africa and central/southern Asia.^1^ In low- and middle-income countries (LMICs), home-based postnatal care interventions, including community health worker (CHW) home visit assessments and referrals of sick newborns, have been shown to reduce neonatal mortality.^2, 3^ Infections are a leading cause of neonatal mortality, with sepsis accounting for 20% of neonatal deaths.^4^ More than half of sepsis-related neonatal deaths in LMICs occur after the first week of age.^5^ Therefore, home-based identification of life-threatening illnesses after the first week of age and referral to hospital are critical to reducing infant morbidity and mortality in LMICs.

The World Health Organization (WHO) recommends assessment of eight clinical signs (‘WHO danger signs’) by first-level health workers, including CHWs, during each postnatal home visit to promote timely identification of sick young infants (<2 months): not feeding well, history of convulsions, fast breathing (>60 breaths per minute), severe chest in-drawing, no spontaneous movement, fever (≥37.5°C), low body temperature (<35.5°C), and any jaundice in the first 24 hours of life or yellow palms and soles at any age.^6, 7^ Identification of any one sign indicates need for referral for further evaluation.^7^

In a recent secondary analysis of a birth cohort in Bangladesh, India, and Pakistan (ANISA), each of seven signs of possible serious bacterial infection (pSBI; seven of the eight WHO danger signs, excluding jaundice) assessed by CHWs in young infants during scheduled home visits was significantly associated with mortality.^8^ However, relying solely on WHO danger signs during routine home visits presents potential challenges. First, WHO danger signs were originally derived among infants brought to health facilities due to caregiver concern.^9^ These infants likely had higher pretest probability of requiring hospital-based care than infants assessed during routine home visits. Second, external validation of the WHO eight danger signs algorithm in a routine home visit setting^10^ was limited to a small sample size (n=395) during the first week of age. Third, the algorithm had high specificity (95%) but lower sensitivity (69%) for physician-assessed severe illness requiring referral, raising concern that its use may miss cases.^10^ Fourth, although WHO guidelines recommend multiple CHW home visits, the current algorithm considers the data from each visit in isolation, and does not utilize accumulated information. The WHO danger signs are employed as *visit-specific* time-varying predictors (e.g., occurrence of fever (yes/no) at each visit). However, beyond the infant’s most recent visit, it is also possible to summarize repeated assessments of clinical signs at prior visits as *aggregative* time-varying predictors (e.g., cumulative number of visits with fever). Aggregative time-varying predictors may better reflect illness trajectory and improve predictive performance compared to a model based on the WHO danger signs, while acknowledging that aggregative time-varying predictors depend on repeated assessments and retention of data from prior visits which may affect feasibility. Lastly, clinical sign-based algorithms, including the WHO danger signs algorithm, to predict sepsis and death in young infants have largely been developed using traditional regression.^11, 12^ Machine learning models such as random forest have been shown to outperform traditional regression approaches to identify serious bacterial infections in febrile young infants presenting to emergency departments in a high-income country.^13^ A recent scoping review of clinical prediction models to diagnose neonatal sepsis in LMICs identified some machine learning models.^14^ However, these models rely on laboratory test predictors and were developed for use in neonatal intensive care units. To our knowledge, machine learning models to predict severe illnesses and death in infants using clinical features assessed by CHWs during routine home visits remain unexplored. Advantages of machine learning models such as random forest over traditional regression approaches is that they can flexibly model non-linear relationships between predictors and an outcome, naturally account for interactions among predictors, and handle a larger number of predictors without being prone to overfitting.^15^

We hypothesized that the predictive performance of the WHO danger signs algorithm could be improved in two ways: first, by incorporating aggregative time-varying predictors that summarize repeated assessments across home visits; and second, by applying a machine learning approach such as random forest. Therefore, using two modeling methods – time-varying Cox regression and random forest – each applied to the same set of CHW-assessed candidate clinical features, including aggregative time-varying features, we aimed to develop a prediction model for hospitalization and/or death among young infants during routine home visits in Dhaka, Bangladesh.

## METHODS

### Study design and data source

This was a secondary analysis of data from the Synbiotics for the Early Prevention of Severe Infections in Infants (SEPSiS) observational cohort study (NCT04012190).^16^ From November 25, 2020 to February 18, 2022, mother-infant pairs were screened for eligibility at two government healthcare facilities in Dhaka: Maternal and Child Health Training Institute (Azimpur) and Mohammadpur Fertility Services and Training Centre. Infants born generally healthy were enrolled between day 0 (birth) and day 4 of age. This cohort (n=1939) has been described separately^17^ and detailed inclusion/exclusion criteria are in **Supplemental Panel S1**. Community health research workers (CHRWs) conducted infant assessments at up to 11 scheduled in-person home visits at 3- and 6-days post-enrolment, and on days 10, 14, 21, 28, 35, 42, 49, 56, and 60 postnatal age. If in-person visits were not feasible, assessments were attempted by telephone. Infants with at least one in-person CHRW home visit after enrolment were included in this analysis. The observation period was from the first CHRW home visit to day 67 of age, allowing predictors ascertained up to the 60-day visit to predict events up to day 67.

This secondary-use study was approved by The Hospital for Sick Children Research Ethics Board (REB #1000079158) and followed the TRIPOD+AI (Transparent Reporting of a multivariable prediction model for Individual Prognosis or Diagnosis+Artificial Intelligence) guidelines.^18^ Caregivers/public were not involved in study design, conduct, or dissemination.

### Outcome

The outcome was time to an event, defined as either first hospitalization and/or death, following the first CHRW visit up to day 67 of age. If an infant was hospitalized and subsequently died, this counted as one event at time of hospitalization. If an infant had multiple hospitalizations, only the first hospitalization was included. Hospitalizations recommended by physicians but declined by caregivers were included. Hospitalizations prior to the first CHRW visit were excluded but these infants were included in the analysis following that visit. Two infants died before their first CHRW in-person visit and were excluded. Hospitalizations for elective surgeries or trauma were excluded. Physicians recommending hospitalization were not study staff, but they may have been informed by study physicians about CHRW findings.

### Primary predictors

Primary candidate predictors were 45 clinical features (25 symptoms from caregiver history, 20 physical exam signs) assessed by CHRWs during scheduled home visits. The primary analysis only included clinical features from in-person visits. Using a standardized checklist, trained CHRWs assessed symptoms and signs occurring on the scheduled visit day, in the last 7 days, or since the last study visit (whichever was most recent). Each of the 45 clinical features was operationalized as *visit-specific* (i.e., based on data obtained at a single visit) and *aggregative* (i.e., based on accumulated data from prior visits) time-varying predictors (**Supplemental Tables S1, S2, S3**). Candidate predictors included the WHO eight danger signs (which, as used in the WHO danger signs algorithm, are visit-specific). Per the SEPSiS protocol, CHRW ascertainment of any WHO danger sign mandated referral to a study physician.

### Additional covariates

Additional covariates were included as predictors based on clinical judgment. Time-fixed additional covariates obtained at enrolment or the first scheduled visit included maternal age, maternal education, antenatal care, infant sex, gestational age at birth, preterm birth (<37 weeks), birth weight-for-gestational age Z-score, umbilical cord care, and any perinatal/delivery complication. Time-varying additional covariates included maternal postpartum substance use, exclusive breastfeeding status, and systemic antibiotic administration on the visit day, in the last 7 days or since the last visit (whichever was most recent). Details on the ascertainment and derivation of additional covariates can be found in **Supplemental Panel S2**.

### Sample size and missing data

Sample size was fixed based on available data from the SEPSiS study eligible for this secondary analysis. Missing values of clinical features were planned to be imputed, if necessary, using multiple imputation.^19^ However, given minimal missingness of clinical features (1.2% to 1.4%), models were developed using complete case analysis.

### Statistical analysis

Demographic characteristics were described using frequencies and percentages for categorical variables, and continuous variables were summarized by means and standard deviations, or medians and interquartile ranges.

#### Variable selection using unadjusted analyses

Variables were selected for inclusion in prediction models based on clinical and statistical factors. Unadjusted time-varying Cox regression analyses were performed to estimate hazard ratios (HRs) with 95% confidence intervals (CI) and p-values. Prevalence of each feature was also considered during predictor selection. In the primary analysis for the time-varying Cox model, clinical features with prevalence ≥1% were used. In the primary analysis for the random forest model, which can handle a larger number of predictors than a regression model while avoiding overfitting, clinical features with prevalence ≥0.1% (instead of 1%) were used. For both the time-varying Cox model and the random forest model, among all aggregative and visit-specific operationalizations for each candidate clinical feature (**Supplemental Tables S1, S2, S3**), the operationalization with the lowest p-value <0.2 in unadjusted analyses^20^ was selected for multivariable analysis. Multicollinearity was assessed using the variance inflation factor (VIF) and variables with the highest VIFs ≥5 were sequentially removed until all VIFs were <5.^21^

#### Time-varying Cox model

Using predictors selected based on unadjusted analyses and multicollinearity, backward selection with a threshold p-value of <0.2 was used in a time-varying multivariable Cox regression analysis to derive a prediction model.^20^ After backward selection, additional covariates and clinical features with p-value <0.2 were re-included in the model if this improved the C-statistic, or removed if this did not change the C-statistic, to achieve the most accurate and parsimonious model. Internal validation was done using 5-fold cross validation.^20^

Additional analyses included: 1) evaluating discrimination and calibration of a time-varying Cox model based on the WHO eight danger signs and a time-varying Cox model based on a single predictor denoting at least one danger sign; 2) adding a single predictor denoting at least one WHO danger sign to the best-performing Cox model in the primary analysis; 3) using only additional covariates and clinical features from caregiver history (excluding physical exam signs); 4) removing hospitalizations with a primary admission diagnosis of jaundice; 5) using a prevalence threshold of ≥0.1% instead of ≥1% for predictor inclusion in multivariable analysis; and 6) after backward selection, not re-including or removing additional covariates with p-value <0.2 from models if this improved discrimination. The rationale for the analysis excluding jaundice hospitalizations is that we observed a high proportion of hospitalizations for jaundice and a strong association between the jaundice predictor and these hospitalizations. We sought to evaluate whether model performance was retained for other clinically significant illnesses.

Sensitivity analyses included: 1) using a threshold p-value of <0.05 instead of <0.2 for backward selection; 2) excluding infants who ever had systemic antibiotic administration during routine home visit assessments; 3) including events up to four weeks after the last CHRW visit; 4) excluding hospitalization events <48 hours duration; 5) imputing missingness of clinical features with last observation carried forward; and 6) reducing the maximum number of scheduled visits.

Subgroup analyses included assessing discrimination of the best-performing model by 1) age group (0-28 days versus 28-60 days), 2) sex, and 3) maternal education level.

#### Random forest model

Using the set of predictors selected based on unadjusted analyses and multicollinearity, we derived a random forest model that emulated the Random Forest for Survival, Longitudinal, and Multivariate data analysis (RF-SLAM) model developed by Wongvibulsin et al. that can handle repeated measurements with time-varying predictors.^22^ The RF-SLAM model was a collection of 1000 decision trees.^22, 23^ The number of features to try at each potential split was the square root of the number of predictors in the model. Each tree was developed from bootstrap samples (100 iterations). Individual infants were bootstrapped rather than bootstrapping infant-days to preserve the original data structure. Randomly selected predictors were used as candidate features to split each node. A Poisson regression log-likelihood was used as the split criterion since it accounted for the division of the follow-up time into infant-days and did not impose the proportional hazards assumption that the predictors have a common effect across the entire follow-up time.^22^ The tree was grown under the constraint that the minimum terminal node size was at least 10% of the total number of infant-days to retain heterogeneity in the terminal nodes.^22^ Variable importance of features was evaluated and compared using the minimal depth, a measure of the predictiveness of a variable in a decision tree algorithm.^24^ Missing data was imputed during the tree growing process by randomly drawing from the non-missing in-bag data within the current node.^22, 23^

In an additional analysis, predictors were selected by applying the same methods outlined above but using a prevalence threshold of ≥1%, instead of ≥0.1%, for selection for inclusion in the multivariable analysis. A further analysis applied the RF-SLAM approach to the predictors included in the best-performing time-varying Cox model.

The performance of all models was evaluated using discrimination (C-statistic and time-dependent area under the receiver operating characteristic curve (AUC) with 95% CIs generated by bootstrapping) and calibration (visual assessment of calibration plots).^25^

All analyses were done using R version 4.4.0 software.^26^

## RESULTS

A total of 1906 infants had at least one CHRW home visit and were included (**Figure 1**). Of these, 176 had a hospitalization and/or death event (**Figure 1**). Admission diagnoses and causes of death are shown in **Supplemental Tables S4 and S5**. CHRWs conducted 20,472 visits (82% in-person, 18% by telephone). The median (25^th^, 75^th^) number of visits (in-person and telephone) per infant was 11 (10, 11). Sixty-two percent of events occurred during the first four weeks of age and the median (25^th^, 75^th^) interval between an event and the most recent CHRW visit was 2 (1, 5) days (**Supplemental Figures S1 and S2**). Of the additional covariates (**Table 1**), gestational age, any perinatal/delivery complication, umbilical cord care, and exclusive breastfeeding status and systemic antibiotic administration since the last visit met the p-value <0.2 threshold and were included in multivariable analyses.

**Figure 1.**
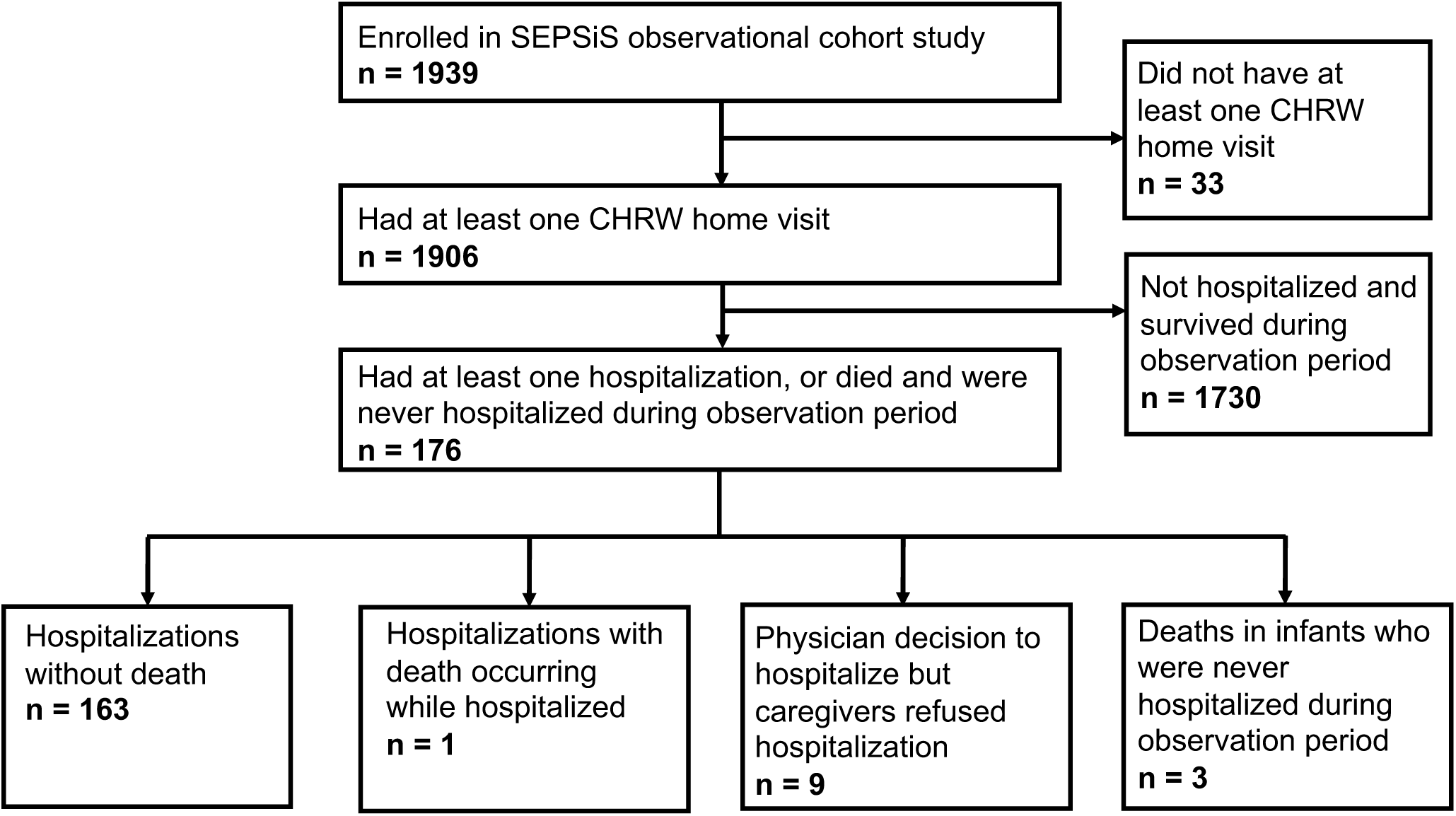
Study flow diagram.

**Table 1.**
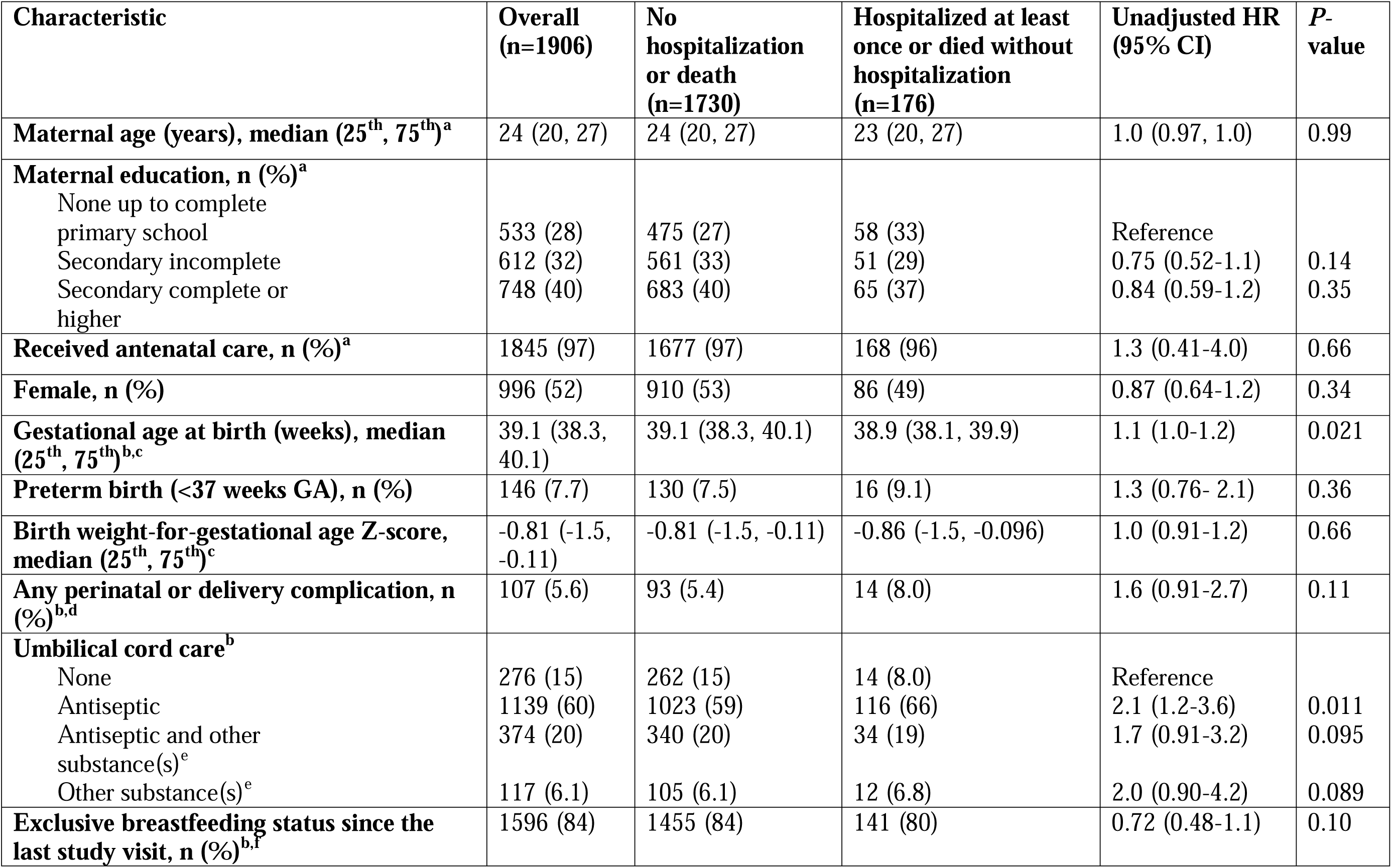
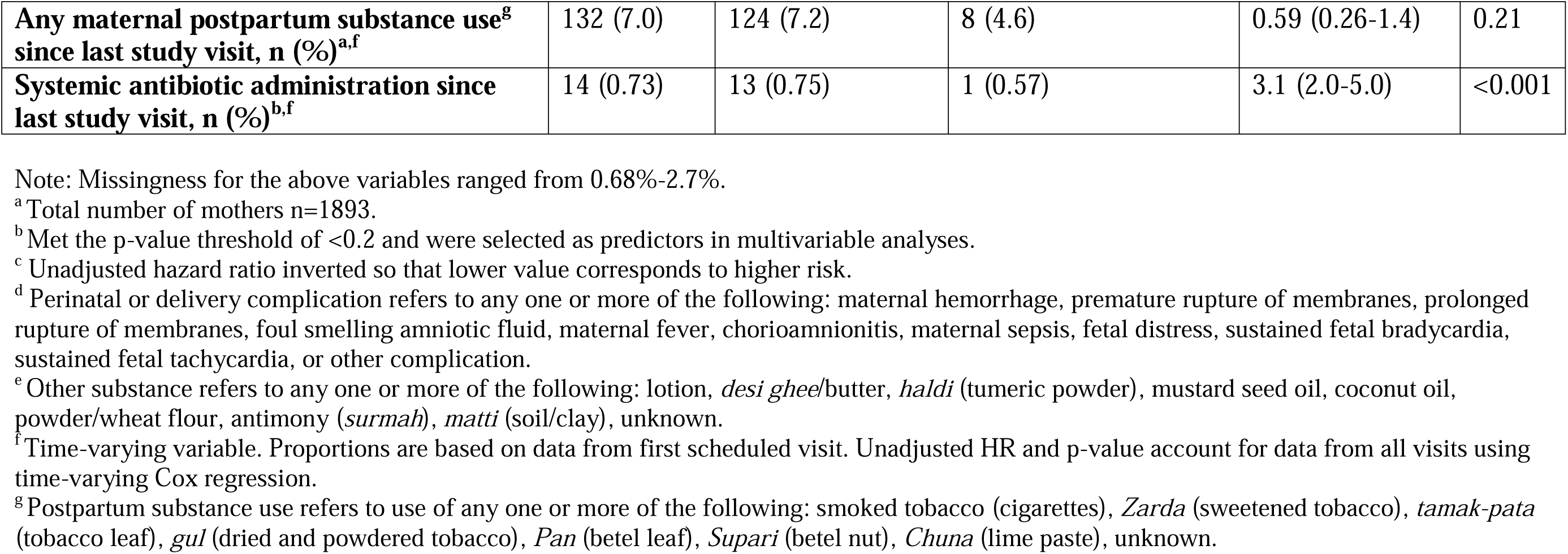
Baseline characteristics and additional covariates and their associations with hospitalization and/or death in unadjusted analyses

Among the clinical features assessed by CHRWs, predictors selected using the p-value from unadjusted analyses and multicollinearity assessment for the primary analysis (lowest p-value <0.2 in unadjusted analysis, VIF<5, prevalence ≥1%) were all visit-specific time-varying operationalizations except for the cumulative sum of visits with red, oozing and/or swollen eyes up to the last visit, which was an aggregative time-varying predictor (**Supplemental Table S6**). All other aggregative time-varying operationalizations had higher p-values, and hazard ratios attenuated to the null, compared with corresponding visit-specific variables, and were therefore not carried forward to multivariable analysis (**Supplemental Table S7**).

The best-performing multivariable Cox model included seven predictors and had a C-statistic of 0.71 (95% CI 0.68-0.75) with a similar cross-validated C-statistic (**Table 2**). The model had a consistent time-dependent AUC across the observation period (**Figure 2**).

**Figure 2.**
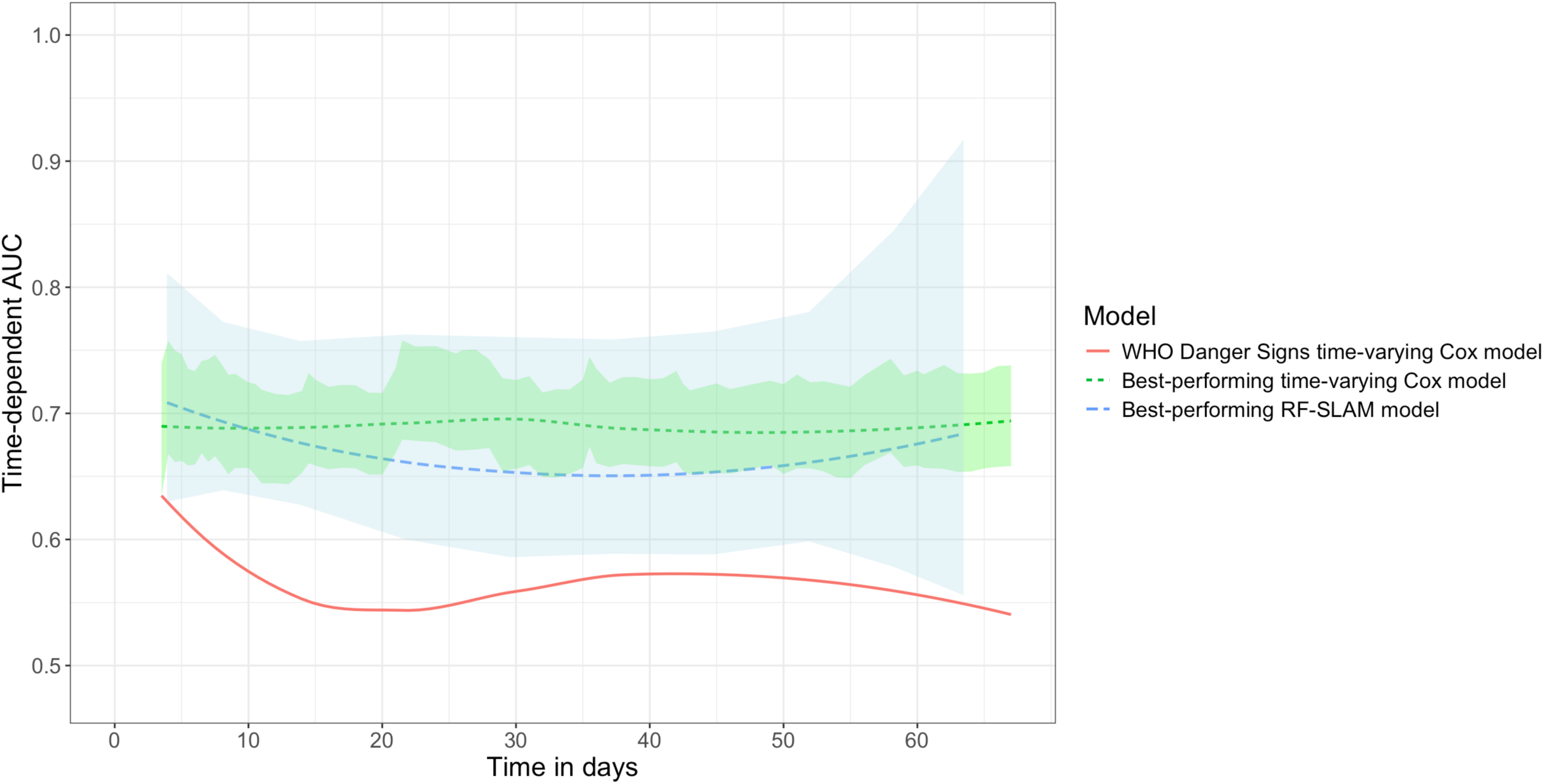
Time-dependent area under the receiver operating characteristic curve (AUC) for best-performing time-varying Cox model for hospitalization and/or death among young infants (using prevalence of predictors ≥1%), time-varying Cox model based on World Health Organization danger signs, and best-performing RF-SLAM model. AUC: Area under the receiver operating characteristic curve; WHO: World Health Organization. Note: Green and blue shaded areas represent 95% confidence intervals (CIs). The 95% CI for the WHO danger signs time-varying Cox model (red) is not shown because the CI was very wide and not useful for illustrative purposes.

**Table 2.** Discrimination of best-performing time-varying Cox model; time-varying Cox model based on World Health Organization (WHO) danger signs; and best-performing time-varying Cox model limited to clinical features (with additional covariates removed) plus a variable denoting at least one WHO danger sign for hospitalization and/or death among young infants (using predictors with prevalence ≥1%)

|  | Predictor | Prevalence, n (%)<sup>a</sup> | Adjusted HR (95% CI) | P-value | C-statistic (95% CI) | Cross-validated C-statistic (95% CI) |
| --- | --- | --- | --- | --- | --- | --- |
| **Best-performing time-varying Cox model** |  |  |  |  |  |  |
| **Additional covariates** | Any perinatal or delivery complication | 864 (5.4) | 1.3 (0.71-2.2) | 0.43 | 0.71 (0.68-0.75) | 0.68 (0.58-0.78) |
|  | Umbilical cord care |  |  |  |  |  |
|  | Antiseptic | 9546 (60) | 1.6 (0.92-2.8) | 0.096 |  |  |
|  | Antiseptic and other substance(s) | 3241 (20) | 1.5 (0.79-2.8) | 0.23 |  |  |
|  |  | 941 (5.9) |  |  |  |  |
|  | Other substance(s) |  | 1.6 (0.71-3.5) | 0.26 |  |  |
|  | Gestational age at birth (weeks), median (25<sup>th</sup>, 75<sup>th</sup>)<sup>b</sup> | 39.1 (38.3, 40.1) | 1.1 (1.0-1.2) | 0.032 |  |  |
| History | Stuffy nose | 1142 (7.1) | 3.0 (2.0-4.6) | <0.001 |  |  |
|  | Yellow discoloration of skin or eyes | 199 (1.2) | 11 (6.6-17) | <0.001 |  |  |
|  | Unusual skin rash or anything abnormal on skin | 183 (1.1) | 2.3 (1.1-4.8) | 0.024 |  |  |
| Physical exam | Cough | 178 (1.1) | 7.5 (4.4-13) | <0.001 |  |  |
| Time-varying Cox model based on World Health Organization danger signs |  |  |  |  |  |  |
| History | Abnormal movement (convulsions/fits) | 9 (0.056) | 15 (2.1-107) | 0.0073 | 0.56 (0.54-0.60) | 0.51 (0.47-0.55) |
| Physical exam | Not sucking effectively | 17 (0.11) | 14 (4.2-46) | <0.001 |  |  |
|  | Fast breathing (RR >60 per minute) | 19 (0.12) | 2.3 (0.48-12) | 0.30 |  |  |
|  | Severe lower chest wall in-drawing | 14 (0.087) | 20 (5.0-81) | <0.001 |  |  |
|  | Lethargy (no movement or movement only on stimulation) | 4 (0.025) | 48 (8.4-275) | <0.001 |  |  |
|  | Fever (one temperature measurement ≥38°C or two measurements ≥37.5°C)<sup>c</sup> | 11 (0.069) | 32 (9.9-102) | <0.001 |  |  |
|  | Low body temperature (one temperature measurement <34.5°C or two measurements<br><35.5°C)<sup>c</sup> | 7 (0.047) | 2.4 (0.13-43) | 0.56 |  |  |
|  | Yellow<br>discoloration of<br>hands and soles<br>of feet | 13 (0.081) | 31 (12-78) | <0.001 |  |  |
| **Physical**<br>**exam** | 1 of any of the 8<br>danger signs | 83 (0.52) | 20 (12-33) | <0.001 | 0.56 (0.53-0.60) | 0.54 (0.49-0.58) |
| **Best-performing time-varying Cox model limited to clinical features (with additional covariates removed) plus a variable denoting at least one World Health Organization danger sign** |  |  |  |  |  |  |
| **History** | Stuffy nose | 1142 (7.1) | 3.0 (2.0-4.5) | <0.001 | 0.70 (0.67-0.74) | 0.68 (0.59-0.76) |
|  | Yellow<br>discoloration of<br>skin or eyes | 199 (1.2) | 8.9 (5.5-14) | <0.001 |  |  |
|  | Unusual skin<br>rash or anything<br>abnormal on skin | 183 (1.1) | 2.9 (1.4-5.9) | 0.0044 |  |  |
| **Physical**<br>**exam** | Cough | 178 (1.1) | 5.6 (3.1-10) | <0.001 |  |  |
|  | 1 of any of the 8<br>WHO danger<br>signs | 83 (0.52) | 6.0 (3.4-11) | <0.001 |  |  |

<sup>a</sup> Denominator is total number of in-person visits (n=16,023).
<sup>b</sup> Adjusted hazard ratio inverted so that lower gestational age corresponds to higher risk.
<sup>c</sup> Thresholds for fever and low body temperature vary slightly across WHO guidelines.
Note: The cumulative sum of visits with red, oozing and/or swollen eyes up to the last visit prior to the current visit met the criteria to be selected as a candidate predictor and was included after backward selection (**Supplemental Table S6**). Since this predictor was the only aggregative time-varying operationalization in the final set of candidate predictors, its removal would support the feasibility of the final best-performing time-varying Cox model. Removing this predictor changed the C-statistic of the best-performing time-varying Cox model minimally from 0.72 (95% CI 0.69-0.76) to 0.71 (95% CI 0.68-0.75) and it was therefore removed to achieve a more parsimonious and feasible final multivariable model.

The WHO eight danger signs were infrequent (<0.1% of visits). A time-varying Cox model based on WHO danger signs or a single variable denoting at least one of these signs both had a C-statistic of 0.56 (95% CI 0.54-0.60) with a similar cross-validated C-statistic (**Table 2**). The time-dependent AUC value of the WHO danger signs was higher during the first week of age and subsequently decreased (**Figure 2**).

The best-performing time-varying Cox model with the addition of a variable denoting at least one WHO danger sign had a C-statistic of 0.73 (95% CI 0.69-0.76) (**Supplemental Table S8**). A time-varying Cox model consisting of the four clinical features from the best-performing Cox model plus a variable denoting at least one WHO danger sign had a C-statistic of 0.70 (95% CI 0.67-0.74) (**Table 2**).

In an additional analysis limiting predictors to baseline additional covariates and caregiver-reported symptoms on history alone during in-person visits, the best-performing time-varying Cox model discrimination was similar (**Supplemental Table S9**). When the additional covariates in the best-performing time-varying Cox model were removed, leaving only four clinical features, the C-statistic decreased slightly (**Supplemental Table S10**).

Other additional, sensitivity, and subgroup analyses demonstrated no substantial changes in discrimination (**Supplemental Tables S11, S12, S13, S14**).

The best-performing time-varying Cox model (**Table 2**) was derived using data from up to 11 CHRW visits. As the maximum number of CHRW home visits per infant decreases, thereby increasing the average prediction window (i.e., number of days between an event and the most recent visit), the C-statistic decreases (**Supplemental Figure S3**).

Across all deciles of predicted risks, both the best-performing time-varying Cox model and WHO danger signs Cox model overestimated predicted risks with gradually increasing differences between predicted and observed hazard rates and 95% CIs excluding 0 (**Figure 3**).

**Figure 3.**
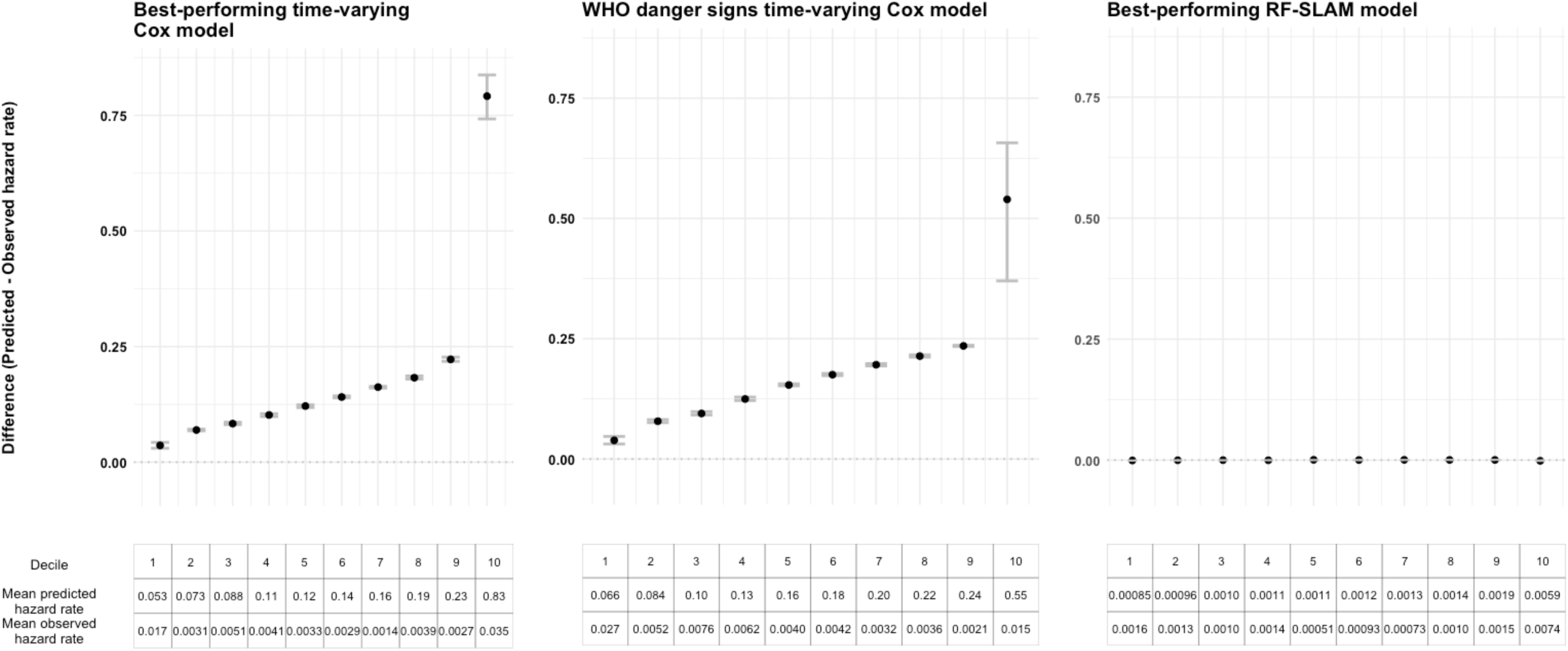
Calibration plots by decile of predicted hazard rates for best-performing time-varying Cox model, time-varying Cox model based on World Health Organization danger signs, and best-performing RF-SLAM model Discrete risk groups were defined by deciles of the predicted hazard rates with the first decile having the lowest predicted hazard rates and the 10^th^ decile having the highest predicted hazard rates. Calibration was assessed by risk group. For each decile risk group, the mean predicted hazard rate was compared with the mean observed hazard rate. The difference between the mean predicted and observed hazard rates was computed and then plotted for each corresponding decile. The gray bars indicate the 95% confidence intervals from 1000 bootstrapped data sets. For well-calibrated models, the difference between the predicted and observed hazard rates should be minimal (near 0).^25^

The 42 predictors in the best-performing RF-SLAM model (p-value <0.2 in unadjusted analysis, VIF<5 and prevalence ≥0.1%), which included both visit-specific and aggregative predictors, are shown in **Supplemental Figure S4**. Variables with predictive importance are those with a minimal depth less than the depth threshold of 10.99. The best-performing RF-SLAM model had a C-statistic of 0.69 (95% CI 0.64-0.73) and was well-calibrated (**Figure 3**). The RF-SLAM model had a higher AUC at the start and end of the observation period (**Figure 2**).

A RF-SLAM model using predictors with a prevalence of ≥1%, instead of ≥0.1% (13 predictors, all baseline covariates or visit-specific clinical features), had a C-statistic of 0.64 (95% CI 0.61-0.69) (variable importance plot shown in **Supplemental Figure S5**).

A RF-SLAM model using the seven predictors (three baseline covariates and four visit-specific clinical features) included in the best-performing time-varying Cox model had a C-statistic of 0.66 (95% CI 0.63-0.72) (variable importance plot shown in **Supplemental Figure S6**).

## DISCUSSION

A Cox model using time-fixed baseline covariates and visit-specific time-varying clinical features demonstrated higher discrimination and similar calibration compared to the conventional approach using WHO danger signs to predict hospitalization and/or death among young infants during CHW routine home visits in Dhaka, Bangladesh. Discrimination was maintained using four clinical features from the best-performing Cox model (nasal congestion, jaundice, skin rash, cough) plus a variable denoting at least one WHO danger sign. Aggregative time-varying operationalizations of predictors were less associated with hospitalization and/or death than visit-specific operationalizations, were not retained in the best-performing Cox model, and did not add substantial discriminatory value in the best-performing RF-SLAM model. The best-performing RF-SLAM model was well-calibrated with similar discrimination to the best-performing time-varying Cox model.

In a 2024 systematic review of the diagnostic accuracy of clinical sign algorithms to identify sepsis in young infants,^11^ the most accurate algorithm in the routine home visit setting was the WHO danger signs. None of the included studies developed time-varying models. Although WHO danger signs had poorer discrimination than the best-performing Cox model in the present secondary analysis of the SEPSiS cohort, they remained important given their high specificity (∼95%) for severe illness requiring referral^10^ and significant association with mortality.^8^ However, in this SEPSiS cohort of infants born generally healthy in an urban setting, WHO danger signs were infrequent at routine home visits (<0.1% of visits). In contrast, in ANISA, a large multi-country south Asian birth cohort including rural areas, prevalence of at least one sign of pSBI during CHW scheduled home visits was substantially higher at 2.7%.^8^ This lower prevalence of WHO danger signs in SEPSiS compared to ANISA may be due to SEPSiS excluding higher-risk infants, including infants with very low birthweight (<1500g), major congenital gastrointestinal anomalies, maternal HIV exposure, need for mechanical ventilation and/or cardiac support (e.g., inotropes), receipt of parenteral antibiotics, or high likelihood of death or major surgery within the first week of age. In contrast, ANISA had no clinical exclusion criteria.^8^ Furthermore, infants in SEPSiS were recruited at government health facilities which may have led to the considerable differences in baseline characteristics compared to infants in ANISA,^8^ 54% of whom were born at a health facility and 46% of whom were born at home. Compared to ANISA,^8^ SEPSiS infants had substantially higher maternal antenatal care (97% in SEPSiS vs. 39% in ANISA), and much lower proportions of perinatal/delivery complications (5.6% in SEPSiS vs. 16% in ANISA), preterm (<37 weeks gestational age) birth (7.7% in SEPSiS vs. 20% in ANISA), low birthweight (<2500g) (17% in SEPSiS vs. 27% in ANISA), and death (0.26% in SEPSiS vs. 2.7% in ANISA).

Clinical features in the best-performing Cox model, including nasal congestion, jaundice, skin rash, and cough, were much more frequently ascertained during routine visits than WHO danger signs. Forty-two percent of hospitalizations were attributed to pneumonia, many of which were likely viral in etiology; in these cases, milder and more common symptoms such as nasal congestion and cough may have higher predictive accuracy than WHO danger signs. Furthermore, discrimination improved when combining the best-performing Cox model, or its four clinical features, with a variable denoting at least one WHO danger sign compared to danger signs alone. Therefore, WHO danger signs remain essential but may be insufficient to accurately identify the full spectrum of illnesses requiring hospitalization in infants born generally healthy in an urban setting. Adding the four clinical features identified in the best-performing Cox model to the WHO danger signs algorithm may improve its predictive performance.

We hypothesized that aggregative time-varying predictors would have higher predictive accuracy than their corresponding visit-specific operationalizations. From a clinical standpoint, cumulative days or weeks of cough would be more concerning than cough on a single visit. Although there were up to 11 home visits scheduled (median of 11 visits attended), this frequency may not have been sufficient to capture the rapid progression of severe illnesses requiring hospitalization in young infants. For example, if a pneumonia evolved over the course of days 38 to 42 of age, the infant may not have exhibited cough or other concerning symptoms and signs during the recent scheduled preceding visits on days 28 and 35, making the predictor ‘cumulative number of visits with cough’ uninformative. Severe illnesses such as sepsis also typically progress within hours to days, making it unlikely that aggregating data across visits spaced four to seven days apart would provide additional benefit beyond the information available from the most recent visit alone.

We further hypothesized that a machine learning model such as RF-SLAM would outperform the time-varying Cox model. In theory, RF-SLAM has several advantages over traditional regression. First, RF-SLAM is an ensemble tree method that utilizes model-averaging.^22^ This lessens the variation associated with individual trees and should result in more accurate predictions. Second, RF-SLAM may more flexibly model non-linear relationships between predictors and an outcome. In the best-performing RF-SLAM model, axillary temperature and gestational age were the first and third most important predictors, respectively. While gestational age at birth was retained in the best-performing Cox model, axillary temperature was not. For both predictors, values above and below the normal ranges indicate high risk of a poor outcome such as hospitalization or death. The inclusion of axillary temperature as the most important variable in the RF-SLAM model and its exclusion in the Cox model may reflect RF-SLAM’s ability to capture non-linear relationships. Conversely, for the Cox model, neither axillary temperature nor dichotomized variables for axillary temperature using prespecified thresholds (i.e., ‘Fever ≥37.5°C’ and ‘Low body temperature <35.5°C’) were retained in the best-performing model. Third, RF-SLAM can handle more predictors, including those with low prevalence, without being prone to overfitting.^22^ Fourth, tree-based models naturally account for interaction effects because each split in the tree is conditioned on previous splits.^27^ However, tree-based models also have disadvantages. Compared to traditional regression, random forest models such as RF-SLAM are computationally intensive (3-4 hour run time in this study) and have limited capacity to provide clinically interpretable associations between individual covariates and an outcome.

The best-performing RF-SLAM model included 42 predictors compared with seven predictors in the best-performing Cox model. However, even when including a larger set of predictors, and despite the theoretical advantages of random forest, the best-performing RF-SLAM model had similar discrimination as the best-performing Cox model. Compared to the best-performing Cox model, the best-performing RF-SLAM model demonstrated better calibration. When the RF-SLAM model was restricted to the seven predictors included in the best-performing Cox model, discrimination was slightly lower, but comparable to that of the best-performing Cox model. These findings suggest that using machine learning models such as random forest may not always result in higher discrimination. While some studies have demonstrated that machine learning methods, including random forest, can improve discrimination compared to traditional regression,^13, 28^ another study found that regression performed comparably to machine learning models.^29^ The improved calibration of the RF-SLAM model compared with the time-varying Cox model may be due in part to its Poisson log-likelihood split function, which does not rely on the proportional hazards assumption and can naturally accommodate time-varying effects. This flexibility may allow RF-SLAM to better match observed hazard rates across risk strata.^22^ Furthermore, because RF-SLAM predictions are averaged across many trees,^22^ the model is less affected by extreme or sparse predictor combinations, which may also contribute to its superior calibration relative to the time-varying Cox model.

A limitation of this study was that the observation period started at the first CHRW home visit (day 3 to 7 of age), thereby excluding the first 24 hours of age when neonatal mortality is highest.^30^ However, infants in this setting remained at high risk of hospitalization and death from days 3 to 67 of age, substantiating the importance of prediction models applicable to this period. Second, prevalence of symptoms and signs during routine home visits was relatively low. In the urban setting of Dhaka where health facilities are numerous and caregivers readily seek care for infant illness, CHWs are less likely to ascertain concerning features during routine home visits. In a separate analysis of severe infection incidence in this cohort, 85% of severe infection episodes were identified following caregiver self-referral compared to 15% identified following a scheduled CHRW home visit assessment and referral.^17^ External validation of the model is needed, particularly in settings with more limited access to health facilities and where caregivers may not as readily seek care for infant illness (e.g., rural communities). Third, the baseline additional covariates in the best-performing Cox model may not be readily available outside of a research setting. For example, information regarding gestational age at birth was obtained from a variety of sources including the mother’s antenatal card and ultrasound report. Nevertheless, a sensitivity analysis removing the baseline additional covariates did not demonstrate a substantial decrease in discrimination. Fourth, the protocol-defined referral system, mandating referral to a study physician if a CHRW ascertained one of the WHO danger signs during a visit, may have increased the association of the danger signs with hospitalization, thereby potentially introducing a bias that may improve discrimination of the WHO danger signs model. However, despite this potential protocol-induced bias, the best-performing Cox model did not include any of the WHO danger signs and showed higher discrimination than the WHO danger signs model. Fifth, although physicians assessing the outcome were not study staff, there was no protocol-instituted blinding to CHRW assessments, and their recommendations for hospitalization could have been influenced by CHRW findings.

### Implications for clinical use and future research

The best-performing Cox model (C-statistic=0.71) may lack sufficient discrimination in its current form to reliably guide CHW referral decisions. Furthermore, this model was derived using up to 11 CHRW home visits and requires baseline data which may be considered too resource-intensive in many settings. A trend of decreasing discrimination was observed as the maximum number of home visits decreased. However, when the maximum number of visits was reduced to four, as recommended by current WHO guidelines,^7^ model discrimination was similar. Therefore, although using up to 11 visits increases model discrimination, this high number of visits may not be essential. When implementing this model, one must consider the trade-off between the labour and costs associated with the number of CHW visits and model discrimination.

If in-person CHW visits are feasible, a combination of the best-performing Cox model plus a variable denoting at least one WHO danger sign achieved the highest discrimination. Considering the potential challenges with ascertaining baseline covariates such as gestational age in LMIC settings, removing the additional covariates from the best-performing Cox model and simplifying it to its four clinical features plus a variable denoting at least one WHO danger sign showed similar discrimination. However, before this simplified model plus WHO danger signs can be implemented into practice, it requires external validation in both urban and rural LMIC settings. Future research should also evaluate whether adding clinical features to the WHO danger signs algorithm can improve identification of infants needing referral across diverse settings and with varying baseline risks. Lastly, backward selection using a threshold p-value is commonly used for predictor selection,^31^ but other methods exist^32–34^ and may be considered in future studies.

## CONCLUSION

During CHW routine home visit assessments of infants born generally healthy in an urban setting, aggregative clinical features do not improve prediction of hospitalization and/or death compared to visit-specific clinical features. Despite theoretical advantages of machine learning models such as random forest, a time-varying Cox model consisting of three baseline covariates and four visit-specific clinical features had similar discrimination and would be more feasible in clinical practice. Among infants born generally healthy, adding four clinical features – nasal congestion, jaundice, skin rash, and cough – to the WHO danger signs algorithm may improve its predictive performance by capturing a broader spectrum of illnesses requiring hospitalization.

## DATA AVAILABILITY STATEMENT

De-identified datasets and code files used in the analyses of this study are publicly available at the Borealis online data repository at https://doi.org/10.5683/SP3/AMBJCL under Custom Dataset Terms. Standard operating procedures (SOPs), including those related to SEPSiS observational cohort study enrolment, consent and data collection procedures, are available at https://doi.org/10.5683/SP3/WKDQYY.

## FOOTNOTES

### Study protocol

The protocol for this secondary analysis study is available upon request.

### Contributors

AF, DGB, SKM, PJG, DHH, PSS, SMAG, SY, SAS, SS, JB, and DER conceptualized the study and developed the methodology. AF, MS, CH, and C-YC managed the data and conducted the data analysis. JB and DER supervised data analysis. AF wrote the initial draft of the manuscript. All authors provided feedback on data analysis and the manuscript. All authors read and approved the final manuscript.

### Patient and public involvement

Caregivers/public were not involved in developing the research question, recruitment, design and conduct of the study, or methods for study result dissemination.

### Ethics statement

This secondary analysis study of the SEPSiS Observational Cohort Study was approved by The Hospital for Sick Children Research Ethics Board (REB #1000079158). The SEPSiS Observational Cohort Study was approved by The Hospital for Sick Children Research Ethics Board (REB #1000063899) and the ethical review committees at the International Centre for Diarrhoeal Disease Research, Bangladesh (icddr,b) (PR-19045) and Bangladesh Shishu Hospital and Institute (formally known as Dhaka Shishu Hospital), the ethical governing body for the Child Health Research Foundation (CHRF) (BICH-ERC-20/02/2019). Informed written consent was obtained from a parent or legal guardian before participant enrolment.

## Acknowledgements

We thank Aline Freitas for contributing some of the statistical code that was used in or adapted for this analysis. We thank Safwan Ahmed and Vishva Shah for their assistance in reviewing references and verifying the accuracy and appropriateness of citations. We also thank all SEPSiS co-investigators and study personnel who contributed to the SEPSiS observational cohort study.

## REFERENCES

1. UNICEF and partners in the United Nations Inter-Agency Group for Child Mortality Estimation. Levels and Trends in Child Mortality Report 2024: Estimates developed by the United Nations Inter-agency Group for Child Mortality Estimation: United Nations Children’s Fund; 2025 [Available from: https://data.unicef.org/resources/levels-and-trends-in-child-mortality-2024/.

2. Tiruneh GT, Shiferaw CB, Worku A. Effectiveness and cost-effectiveness of home-based postpartum care on neonatal mortality and exclusive breastfeeding practice in low-and-middle-income countries: a systematic review and meta-analysis. BMC Pregnancy Childbirth. 2019;19(1):507.

3. Lassi ZS, Kedzior SG, Bhutta ZA. Community-based maternal and newborn educational care packages for improving neonatal health and survival in low- and middle-income countries. Cochrane Database Syst Rev. 2019;2019(11).

4. Rudd KE, Johnson SC, Agesa KM, Shackelford KA, Tsoi D, Kievlan DR, et al. Global, regional, and national sepsis incidence and mortality, 1990-2017: analysis for the Global Burden of Disease Study. Lancet. 2020;395(10219):200-11.

5. Sankar MJ, Natarajan CK, Das RR, Agarwal R, Chandrasekaran A, Paul VK. When do newborns die? A systematic review of timing of overall and cause-specific neonatal deaths in developing countries. J Perinatol. 2016;36 Suppl 1(Suppl 1):S1–s11.

6. World Health Organization. Integrated Management of Childhood Illness: management of the sick young infant aged up to 2 months Geneva: World Health Organization; 2019 [Available from: https://www.who.int/maternal_child_adolescent/documents/management-sick-young-infant-0-2-months/en/.

7. WHO recommendations on maternal and newborn care for a positive postnatal experience Geneva: World Health Organization; 2022 [Available from: https://www.who.int/publications/i/item/9789240045989.

8. Darmstadt GL, Ahmed S, Islam MS, Abdalla S, El Arifeen S, Arvay ML, et al. Association of clinical signs of possible serious bacterial infections identified by community health workers with mortality of young infants in South Asia: a prospective, observational cohort study. EClinicalMedicine. 2025;80:103070.

9. Young Infants Clinical Signs Study Group. Clinical signs that predict severe illness in children under age 2 months: a multicentre study. Lancet. 2008;371(9607):135–42.

10. Darmstadt GL, Baqui AH, Choi Y, Bari S, Rahman SM, Mannan I, et al. Validation of a clinical algorithm to identify neonates with severe illness during routine household visits in rural Bangladesh. Arch Dis Child. 2011;96(12):1140–6.

11. Fung A, Shafiq Y, Driker S, Rees CA, Mediratta RP, Rosenberg R, et al. Diagnostic Accuracy of Clinical Sign Algorithms to Identify Sepsis in Young Infants Aged 0 to 59 Days: A Systematic Review and Meta-analysis. Pediatrics. 2024;154(Suppl 1).

12. Shafiq Y, Fung A, Driker S, Rees CA, Mediratta RP, Rosenberg R, et al. Predictive Accuracy of Infant Clinical Sign Algorithms for Mortality in Young Infants Aged 0 to 59 Days: A Systematic Review. Pediatrics. 2024;154(Suppl 1).

13. Ramgopal S, Horvat CM, Yanamala N, Alpern ER. Machine Learning To Predict Serious Bacterial Infections in Young Febrile Infants. Pediatrics. 2020;146(3).

14. Neal SR, Sturrock SS, Musorowegomo D, Gannon H, Zaman M, Cortina-Borja M, et al. Clinical prediction models to diagnose neonatal sepsis in low-income and middle-income countries: a scoping review. BMJ Glob Health. 2025;10(4).

15. Goldstein BA, Navar AM, Carter RE. Moving beyond regression techniques in cardiovascular risk prediction: applying machine learning to address analytic challenges. Eur Heart J. 2017;38(23):1805–14.

16. SEPSIS Observational Cohort Study in Young Infants in Bangladesh [Available from: https://ClinicalTrials.gov/show/NCT04012190.

17. Fung A, Heasley C, Pell LG, Bassani DG, Shah PS, Morris SK, et al. Severe infection incidence among young infants in Dhaka, Bangladesh: an observational cohort study. BMJ Public Health. 2025;3(2):e002383.

18. Collins GS, Moons KGM, Dhiman P, Riley RD, Beam AL, Van Calster B, et al. TRIPOD+AI statement: updated guidance for reporting clinical prediction models that use regression or machine learning methods. Bmj. 2024;385:e078378.

19. Austin PC, White IR, Lee DS, van Buuren S. Missing Data in Clinical Research: A Tutorial on Multiple Imputation. Can J Cardiol. 2021;37(9):1322–31.

20. Vittinghoff E, Glidden DV, Shiboski SC, McCulloch CE. Regression Methods in Biostatistics. 2nd ed. Boston, MA: Springer; 2012.

21. Lavery MR, Acharya P, Sivo SA, Xu L. Number of predictors and multicollinearity: What are their effects on error and bias in regression? Communications in Statistics - Simulation and Computation. 2019;48(1):27–38.

22. Wongvibulsin S, Wu KC, Zeger SL. Clinical risk prediction with random forests for survival, longitudinal, and multivariate (RF-SLAM) data analysis. BMC Med Res Methodol. 2019;20(1):1.

23. Ishawaran H, Kogalur U, Blackstone E, Lauer M. Random survival forests. Ann Appl Statist. 2008;2(3):841–60.

24. Ishwaran H, Kogalur UB, Gorodeski EZ, Minn AJ, Lauer MS. High-Dimensional Variable Selection for Survival Data. Journal of the American Statistical Association. 2010;105(489):205–17.

25. Steyerberg EW. Chapter 15.3: Calibration. Clinical Prediction Models: A Practical Approach to Development, Validating, and Updating. Statistics for Biology and Health. 2nd ed. Switzerland: Springer; 2019.

26. R Core Team. R: A Language and Environment for Statistical Computing Vienna, Austria.: R Foundation for Statistical Computing; 2025 [Available from: https://www.R-project.org/.

27. Steyerberg EW. Chapter 4.2.11 Advantages and Disadvantages of Tree Models. Clinical Prediction Models: A Practical Approach to Development, Validating, and Updating. Statistics for Biology and Health. 2nd ed. Switzerland: Springer; 2019.

28. Cygu S, Seow H, Dushoff J, Bolker BM. Comparing machine learning approaches to incorporate time-varying covariates in predicting cancer survival time. Sci Rep. 2023;13(1):1370.

29. Spooner A, Chen E, Sowmya A, Sachdev P, Kochan NA, Trollor J, et al. A comparison of machine learning methods for survival analysis of high-dimensional clinical data for dementia prediction. Sci Rep. 2020;10(1):20410.

30. Dol J, Hughes B, Bonet M, Dorey R, Dorling J, Grant A, et al. Timing of neonatal mortality and severe morbidity during the postnatal period: a systematic review. JBI Evid Synth. 2023;21(1):98–199.

31. Moons KG, Kengne AP, Woodward M, Royston P, Vergouwe Y, Altman DG, et al. Risk prediction models: I. Development, internal validation, and assessing the incremental value of a new (bio)marker. Heart. 2012;98(9):683–90.

32. Steyerberg EW. Chapter 11: Selection of Main Effects. Clinical Prediction Models: A Practical Approach to Development, Validating, and Updating. Statistics for Biology and Health. 2nd ed. Switzerland: Springer; 2019. p. 207-25.

33. Austin PC, Tu JV. Bootstrap Methods for Developing Predictive Models. The American Statistician. 2004;58(2):131–7.

34. Tibshirani R. Regression Shrinkage and Selection Via the Lasso. Journal of the Royal Statistical Society: Series B (Methodological). 2018;58(1):267–88.

