## Supplemental File 1 for "Development of a prediction model for infant hospitalization and death using clinical features assessed by community health workers during routine postnatal home visits in Dhaka, Bangladesh"

Alastair Fung^1,2,3,4^, Marimuthu Sappani^5^, Cole Heasley^2^, Chun-Yuan Chen^2^, Shaun K. Morris^1,2,3,4^, Peter J. Gill^1,3,4^, Diego G. Bassani^2,3,4^, Davidson H. Hamer^6,7^, Prakesh S. Shah^1,3,8^, S. M. Abdul Gaffar^9^, Sultana Yeasmin^9^, Shafiqul A. Sarker^9^, Shamima Sultana^9^, Joseph Beyene*^3,5^, Daniel E. Roth*^1,2,3,4^

*Contributed equally as co-senior authors

^1^Department of Paediatrics, The Hospital for Sick Children, University of Toronto, Toronto, Canada.

^2^Centre for Global Child Health, The Hospital for Sick Children, Toronto, Canada.

^3^Dalla Lana School of Public Health, University of Toronto, Toronto, Canada.

^4^Child Health Evaluative Sciences, The Hospital for Sick Children Research Institute, Toronto, Canada.

^5^Department of Health Research Methods, Evidence and Impact, Faculty of Health Sciences, McMaster University, Hamilton, Canada.

^6^Department of Global Health, Boston University School of Public Health, Boston, USA.

^7^Section of Infectious Diseases, Boston University Chobanian and Avedisian School of Medicine, Boston, USA.

^8^Department of Pediatrics, Mount Sinai Hospital and University of Toronto, Toronto, Canada.

^9^International Centre for Diarrhoeal Disease Research, Dhaka, Bangladesh.

**Table of contents**	Page

**Panel S1**. Inclusion and exclusion criteria of SEPSiS observational cohort study	4

**Panel S2**. Ascertainment and derivation of additional covariates by study personnel	6

**Table S1.** Operational definitions of predictor variables	7

**Table S2.** Exposure variables obtained on caregiver history during community health research 8 worker visit and operationalization

**Table S3.** Exposure variables obtained on physical examination during community health	9

research worker visit and operationalization

**Table S4**. Primary admission diagnoses for hospitalizations	14

**Table S5**. Causes of death	15

**Table S6.** Candidate predictors selected and their associations with hospitalization and/or	16

death in unadjusted analyses

**Table S7.** Comparison of associations of visit-specific and aggregative time-varying	17

predictor operationalizations with hospitalization and/or death in unadjusted

analyses (for predictors with prevalence ≥1%)

**Table S8**. Discrimination of best-performing Cox model for hospitalization and/or death	19

among young infants with forced inclusion of World Health Organization danger signs

**Table S9.** Discrimination of best-performing Cox model for hospitalization and/or death	20

among young infants limited to baseline additional covariates plus symptoms on history only

**Table S10.** Discrimination of best-performing Cox model for hospitalization and/or death	21

among young infants limited to clinical features (with additional covariates removed)

**Table S11.** Discrimination of Cox model for hospitalization and/or death among young	22

infants (using prevalence of predictors ≥1%) with removal of jaundice hospitalizations

**Table S12**. Discrimination of Cox model for hospitalization and/or death among young	23

infants (using prevalence of predictors ≥0.1% instead of ≥1%)

**Table S13.** Discrimination of Cox model for hospitalization and/or death among young	24

infants (using prevalence of predictors ≥1%) using only predictors retained after backward selection without manually forcing additional covariates into the model to improve model discrimination

**Table S14.** Sensitivity and subgroup analyses for discrimination of best-performing Cox	25

model for hospitalization and/or death among young infants (using prevalence of predictors ≥1%)

**Figure S1.** Distribution of hospitalizations and deaths in infants who were never hospitalized	26

by week of age

**Figure S2.** Frequency of durations in days between outcome event (hospitalization and/or	27

death) and most recent community health research worker visit (n=176)

**Figure S3**. C-statistic by maximum number of visits per infant for best-performing Cox	28

model for hospitalization and/or death among young infants (using prevalence of

predictors ≥1%)

**Figure S4.** Variable importance by minimal depth for best-performing Random Forest for 29 Survival, Longitudinal, and Multivariate data (RF-SLAM) model for hospitalization and/or
death among young infants (using prevalence of predictors ≥0.1%)

**Figure S5.** Variable importance by minimal depth for Random Forest for Survival, 30

Longitudinal, and Multivariate data (RF-SLAM) model for hospitalization and/or death

among young infants (using prevalence of predictors ≥1% instead of ≥0.1%)

**Figure S6**. Variable importance by minimal depth for Random Forest for Survival, 31

Longitudinal, and Multivariate data (RF-SLAM) model for hospitalization and/or death

among young infants using predictors from best-performing time-varying Cox model

**Panel S1.** Inclusion and exclusion criteria of SEPSiS observational cohort study

*Inclusion criteria*

1. Infants up to and including 4 days of age

2. Infant delivered at a study hospital

3. Orally feeding currently^a^

4. Informed consent by parent or guardian

5. Intends to maintain residence within the defined catchment areas (upon discharge from hospital) until 60 days of age

*Exclusion criteria*

1. Birth weight <1500g^b^

2. Death or major surgery^c^ considered to be highly probable within the first week of age

3. Major congenital anomaly of the gastrointestinal tract

4. Maternal HIV infection and/or history of mother ever receiving anti-retroviral drug(s) for presumed HIV infection^d^

5. Current mechanical ventilation and/or cardiac support (e.g., inotropes) and/or administration/prescription of parenteral antibiotics^a^

6. Any prenatal or postpartum use of non-dietary probiotic supplement by the mother (during current pregnancy)^e^

7. Any postnatal administration of non-dietary probiotic or prebiotic supplements to infant

8. Enrolment of infant in any other clinical trial involving the administration of probiotics and/or prebiotics

9. Resides in the same household as another infant previously enrolled in the study, or any study within the research platform, who is currently <60 days of age; however, twins may be enrolled simultaneously

10. Multiple gestation for which the number of liveborn infants from the same pregnancy exceeds two (i.e., triplets or higher order multiples)

^a^ These criteria were time-varying, so they were reassessed on a daily basis until no longer eligible for another reason (i.e., beyond day 4 of age), as long as the infant remained potentially eligible by other criteria. Orally feeding was defined as being able to take a probiotic or synbiotic supplement by mouth on a daily basis.

^b^ Current infant weight, as measured and documented by study personnel, was used if birth weight was missing, illegibly recorded, or suspected of being an error (e.g., implausible value, discrepancy of greater than 15% between documented birth weight and measured current weight).

^c^ Major surgery was defined as an operative procedure to explore and/or repair an organ or tissue that is performed under general anesthesia. Examples relevant to the neonatal period include ligation of patent ductus arteriosus, repair of abdominal wall defects, repair of bowel perforation due to necrotizing enterocolitis, repair of tracheoesophageal fistula and/or esophageal atresia, and repair of myelomeningocele. Conversely, examples of common procedures in newborns not considered major surgery include circumcision, tongue tie release, and removal of extra digits (polydactyly).

^d^ Although HIV in very young infants was expected to be rare in this context, there was a low but non-zero theoretical risk that a baby born to an HIV-positive mother could be significantly compromised, particularly in cases where in-utero transmission occurred earlier in pregnancy. Both the HIV-positive mother and infant were expected to represent a unique population in regard to their respective microbiomes and excluding them should not affect the generalizability of results to the population as a whole. The number of HIV-positive infants was anticipated to be too low to conduct sub-group analyses, and thus, it would not be possible to make meaningful inferences about this population, even if they were to be included in the study.

^e^ A non-dietary probiotic is a commercial (store-bought) probiotic product that is consumed in the form of a capsule, powder, liquid, etc., although it may be mixed into a food or drink at the time of consumption. In contrast, a dietary probiotic is an ingredient of a food or beverage that either occurs naturally or is added during home production or the commercial manufacturing process (e.g., yogurt or fermented drinks).

**Panel S2.** Ascertainment and derivation of additional covariates by study personnel

All data collected in the SEPSiS observational cohort study were recorded by study personnel using an electronic data capture application run on handheld tablets. On the day of enrolment, trained study nurses obtained data on maternal age, maternal education, antenatal care, and any perinatal/delivery complication via caregiver history. Birth weight was measured by a study nurse or study medical officer in the delivery room or as soon as possible after delivery. During the study nurse encounter on the day of enrolment, information regarding gestational age was obtained from a variety of sources including the mother’s antenatal card and ultrasound report (if available), and gestational age at birth in weeks was estimated following American College of Obstetricians and Gynecologists guidelines. Birth weight-for-gestational age Z-score was then calculated using Intergrowth-21^st^ standards. Infant sex was determined during a baseline physical exam by a study medical officer on the day of enrolment. Information on umbilical cord care (none/antiseptic/other substance(s)/antiseptic plus other substance(s)) was obtained at the first CHRW routine home visit by asking the caregiver if they had applied any topical substances to the umbilical cord of the baby at any time since birth.

Information on additional time-varying covariates (maternal postpartum substance use, exclusive breastfeeding status, and systemic antibiotic administration) was obtained via caregiver history during CHRW routine home visit assessments.

**Table S1.** Operational definitions of predictor variables

| **Time-varying predictor category** | **Operationalization** | **Variable type** | **Definition** | **Observation window** |
| --- | --- | --- | --- | --- |
| **Visit-specific** | **Occurrence** | Dichotomous | Symptom/sign occurred | Within the past 1 week, 2 weeks, and 4 weeks |
| **Aggregative** | **Recurrence** | Continuous | Cumulative number of visits with the symptom/sign | Within the past 2 weeks, 4 weeks, and all preceding weeks since the first community health research worker visit |
|  | **Severity** | Continuous | Minimum, maximum, or mean |  |
|  | **Deviation from prior values** | Categorical | At least 1 and 2 standard deviations above or below the infant’s mean value of the symptom/sign based on prior measurements |  |

**Table S2.** Exposure variables obtained on caregiver history during community health research worker visit and operationalization

|  | **Symptom** | **Measurement** | **Operationalization** |
| --- | --- | --- | --- |
| 1. | Not feeding well | No/Yes/Unknown | Occurrence  Recurrence |
| 2. | Unusually sleepy or could not (or cannot) wake from sleep |  |  |
| 3. | Abnormal movement (convulsions/fits) |  |  |
| 4. | Feels hot to touch or has a fever |  |  |
| 5. | Feels cold to touch or has low body temperature |  |  |
| 6. | Red or discharging umbilicus |  |  |
| 7. | Unusual skin rash or anything abnormal on skin |  |  |
| 8. | Skin pustules or boil |  |  |
| 9. | Cough |  |  |
| 10. | Fast or difficult breathing |  |  |
| 11. | Runny nose |  |  |
| 12. | Stuffy nose |  |  |
| 13. | Not gaining enough weight |  |  |
| 14. | Red, oozing and/or swollen eyes |  |  |
| 15. | Yellow discoloration of skin or eyes |  |  |
| 16. | Diarrhea (described as frequency of stool is too often, the quantity of stool is too much and/or the consistency is too loose compared to the usual) |  |  |
| 17. | Any blood in stool |  |  |
| 18. | Any vomiting |  |  |
| 19. | Any blood in vomit |  |  |
| 20. | Projectile vomiting |  |  |
| 21. | Abdomen is too large and/or swollen |  |  |
| 22. | Drainage from ear |  |  |
| 23. | Sores inside the mouth |  |  |
| 24. | Weak, abnormal or absent cry |  |  |
| 25. | Bouts of unsoothable fretting and crying |  |  |

**Table S3.** Exposure variables obtained on physical examination during community health research worker visit and operationalization

|  | **Sign** | **Measurement** | **Derived exposure definition** | **Operationalization** |
| --- | --- | --- | --- | --- |
| 1. | Respiratory rate^a^ | 1^st^ time: ___ breaths/min  2^nd^ time: ___ breaths/min | **1) Highest Respiratory Rate:** The highest respiratory rate will be used (i.e., only the first measure if the respiratory rate was measured only once or the highest of the two measures if the respiratory rate was measured twice.) | Severity  Deviation above prior values |
|  |  |  | **2) Tachypnea**: ‘Tachypnea’ will be defined as the Highest Respiratory Rate: ≥60 breaths per minute for infants <2 months of age, and ≥50 breaths per minute for infants 2 to 12 months of age per the WHO guidelines on acute respiratory infection in children.^99^ | Occurrence  Recurrence |
| 2. | Level of consciousness and movement | Normal movement/  Movement only on stimulation/ No movement at all or unconscious/Unknown | 1) Movement only on stimulation  2) No movement at all or unconscious | Occurrence  Recurrence |
| 3. | Axillary temperature  (measured twice and, if necessary, a third time)^b^ | 1^st^ time: ______.___^o^C  2^nd^ time:______.___^o^C  3^rd^ time:______.___^o^C | **1) Axillary Temperature:** The mean of the two or three temperature measurements will be used. | Severity  Deviation above prior values  Deviation below prior values |
|  |  |  | **2)** **Fever**:**^c^** A fever was defined as one temperature measurement ≥38^o^C or two or more measurements ≥37.5^o^C.^11^ | Occurrence  Recurrence |
|  |  |  | **3)** **Low body temperature**:**^c^** A low body temperature was defined as one measurement <34.5^o^C or two or more measurements <35.5^o^C.^11^ | Occurrence  Recurrence |
| 4. | Appearance of eyes | No eye abnormalities/Sunken/Red/Oozing/ Swollen/Unknown | 1) Sunken eyes  2) Red eyes  3) Oozing eyes  4) Swollen eyes | Occurrence  Recurrence |
| 5. | Severe lower chest wall in-drawing | No/Yes/Unknown  No/Yes/Unknown | Same as symptom/sign  Same as symptom/sign | Occurrence  Recurrence  Occurrence  Recurrence |
| 6. | Cough |  |  |  |
| 7. | Audible wheeze or whistling breath sounds |  |  |  |
| 8. | Weak, abnormal or absent cry |  |  |  |
| 9. | Nasal discharge/ rhinorrhea |  |  |  |
| 10. | Irritable and inconsolable by parent/ caregiver despite attempted comforting and feeding |  |  |  |
| 11. | Convulsions | No observed convulsion/Observed or suspected convulsion/Unknown | Observed or suspected convulsion | Occurrence  Recurrence |
| 12. | Discharge draining from ear | No/Yes/Unknown  No/Yes/Unknown | Same as symptom/sign  Same as symptom/sign | Occurrence  Recurrence  Occurrence  Recurrence |
| 13. | Ulcers in mouth |  |  |  |
| 14. | Skin pinch | Goes back immediately/Goes back slowly/Not checked | Goes back slowly | Occurrence  Recurrence |
| 15. | Jaundice | Yellow discoloration of skin/Yellow discoloration of hands and soles of feet/Yellow discoloration of eyes/Unknown | 1) Yellow discoloration of skin  2) Yellow discoloration of hands and soles of feet  3) Yellow discoloration of eyes | Occurrence  Recurrence |
| 16. | Skin rash | No/Yes/Unknown | Same as symptom/sign | Occurrence  Recurrence |
| 17. | Skin pustules or abscess | Skin pustules/Skin abscess/Unknown | 1) Skin pustules  2) Skin abscess | Occurrence  Recurrence |
| 18. | Umbilicus red, discolored or discharging pus | No/Yes/Unknown | Same as symptom/sign | Occurrence  Recurrence |
| 19. | Bleeding, bruising or petechiae, or untreated injury | No/Yes/Unknown | Same as symptom/sign | Occurrence  Recurrence |
| 20. | Feeding assessment | Sucking normally/Not sucking effectively/Unable to assess feeding/Unknown | Not sucking effectively | Occurrence  Recurrence |

^a^ Repeat if: ≥60 breaths/min and the infant is ≤60 days of age, or ≥50 breaths/min and the infant is >60 days of age.

^b^ If the temperature is ≥37.5^o^C, unbundle the infant, wait 10 mins and then repeat and record the measurement. If the temperature is <35.5^o^C, wrap the infant in a blanket (or extra clothing), wait 10 minutes and then repeat and record the measurement. If the infant’s temperature is within the normal range (35.5-37.4ºC) it is not necessary to modify the infant’s clothing or bundling prior to taking the repeat temperature. A third temperature measurement will be collected if any two measurements differ by more than plus or minus 1°C, even if both measurements are within the normal range.

^c^ Thresholds for fever and low body temperature vary slightly across WHO guidelines.

**Table S4**. Primary admission diagnoses for hospitalizations

| **Admission diagnosis** | **Frequency** | **%** |
| --- | --- | --- |
| Pneumonia | 73 | 42 |
| Neonatal jaundice | 36 | 21 |
| Suspected sepsis | 27 | 16 |
| Omphalitis | 9 | 5.2 |
| Skin and soft tissue infection | 5 | 2.9 |
| Bronchiolitis | 4 | 2.3 |
| Upper respiratory tract infection | 4 | 2.3 |
| Gastroenteritis | 3 | 1.7 |
| Vomiting | 3 | 1.7 |
| Feeding difficulty | 2 | 1.2 |
| Umbilical granuloma | 2 | 1.2 |
| Seizure | 2 | 1.2 |
| Hematochezia | 1 | 0.6 |
| Intestinal obstruction | 1 | 0.6 |
| Vaginal bleeding | 1 | 0.6 |
| **TOTAL** | **173** | **100** |

**Table S5.** Causes of death

| **Hospitalized prior to death** | **Cause of death** | **Frequency** |
| --- | --- | --- |
| No | Cardiorespiratory failure associated with bacterial infection/septicemia | 1 |
| No | SIDS (sudden infant death syndrome) | 1 |
| No | Severe pneumonia | 1 |
| Yes^a^ | Neonatal sepsis associated with pneumonia. | 1 |

^a^ Counted as an event occurring at the time of hospitalization.

**Table S6.** Candidate predictors selected and their associations with hospitalization and/or death in unadjusted analyses

|  | **Predictor** | **Prevalence, n (%)^a^** | **Missing, n (%)^a^** | **Unadjusted HR (95% CI)** | ***P-*value** |
| --- | --- | --- | --- | --- | --- |
| **History** | Stuffy nose | 1142 (7.1) | 192 (1.2) | 4.9 (3.3-7.3) | <0.001 |
|  | Cough | 691 (4.3) | 192 (1.2) | 5.6 (3.7-8.6) | <0.001 |
|  | Feels hot to touch or has fever within the past 7 days | 508 (3.2) | 0 (0.0) | 2.6 (1.3-5.0) | 0.0047 |
|  | Runny nose | 227 (1.4) | 193 (1.2) | 5.5 (3.0-10) | <0.001 |
|  | Cumulative sum of visits with red, oozing and/or swollen eyes up to the last visit, median (range)^b^ | 0 (0, 8) | 0 (0.0) | 0.48 (0.21-1.1) | 0.081 |
|  | Yellow discoloration of skin or eyes | 199 (1.2) | 193 (1.2) | 12 (7.7-19) | <0.001 |
|  | Unusual skin rash or anything abnormal on skin | 183 (1.1) | 193 (1.2) | 3.9 (1.9-7.9) | <0.001 |
| **Physical Exam** | Axillary temperature (^o^C), median (25^th^, 75^th^)^c^ | 36.3 (36.1, 36.5) | 203 (1.3) | 1.9 (0.88-4.0) | 0.11 |
|  | Cough | 178 (1.1) | 203 (1.3) | 13 (8.1-22) | <0.001 |

Note: Using prevalence of predictors ≥1%, no variance inflation factors were >5.

^a^ Denominator is total number of in-person visits (n=16,023).

^b^ Value for first visit missing by default due to lagged variable and imputed with 0.

^c^ Mean of at least two and up to three temperature measurements during the same visit.

**Table S7.** Comparison of associations of visit-specific and aggregative time-varying predictor operationalizations with hospitalization and/or death in unadjusted analyses (for predictors with prevalence ≥1%)

|  |  | **VISIT-SPECIFIC** | | **AGGREGATIVE** | | | | | |
| --- | --- | --- | --- | --- | --- | --- | --- | --- | --- |
|  |  | **Occurrence at today’s visit, in the past 7 days or since the last visit (whichever was most recent)** | | **Cumulative sum of visits with occurrence of clinical feature across all prior visits up to today’s visit** | | **Cumulative sum of visits with occurrence of clinical feature up to the last visit** | | **Measurement at today’s visit is at least 2 standard deviations above mean measurement across all prior visits** | |
|  | **Predictor** | **Unadjusted HR (95% CI)** | ***P-*value** | **Unadjusted HR (95% CI)** | ***P-*value** | **Unadjusted HR (95% CI)** | ***P-*value** | **Unadjusted HR (95% CI)** | ***P-*value** |
| **History** | Stuffy nose | 4.9 (3.3-7.3) | 1.3e^-15^ | 1.5 (1.3-1.7) | 1.8e^-7^ | 1.2 (0.98-1.5) | 0.074 | - | - |
|  | Cough | 5.6 (3.7-8.6) | 2.9e^-15^ | 1.7 (1.4-2.1) | 1.5e^-7^ | 1.3 (0.91-1.8) | 0.16 | - | - |
|  | Feels hot to touch | 2.6 (1.2-6.0) | 0.021 | 1.3 (0.87-2.1) | 0.19 | 0.95 (0.49-1.8) | 0.87 | - | - |
|  | Runny nose | 5.5 (3.0-10) | 5.1e^-8^ | 2.0 (1.3-2.9) | 0.00064 | 1.2 (0.64-2.1) | 0.60 | - | - |
|  | Red, oozing and/or swollen eyes | 1.9 (0.79-4.8) | 0.15 | 0.79, 0.53-1.2) | 0.27 | 0.48 (0.21-1.1) | 0.081 | - | - |
|  | Yellow discoloration of skin or eyes | 12 (7.7-19) | <2.0e^-16^ | 1.4 (1.3-1.6) | 1.8e^-8^ | 1.3 (0.88-1.9) | 0.18 | - | - |
|  | Unusual skin rash or anything abnormal on skin | 3.9 (1.9-7.9) | 0.00023 | 1.6 (1.2-2.4) | 0.016 | 1.3 (0.73-2.1) | 0.42 | - | - |
| **Physical exam** | Axillary temperature (^o^C) | 1.9 (0.88-4.0) | 0.11 | - | - | - | - | 0.93 (0.50-1.7) | 0.81 |
|  | Cough | 13 (8.1-22) | <2.0e^-16^ | 3.5 (2.4-4.9) | 1.6e^-12^ | 1.8 (0.96-3.5) | 0.065 | - | - |

Note: As described in the Methods, aggregative time-varying predictors such as cumulative number of visits with cough, maximum temperature, and deviation of temperature above prior temperature values were operationalized within various time frames (e.g., past 2 weeks, past 4 weeks, or across all prior visits). The association of each operationalization for each clinical feature listed in **Supplemental** **Table S3** with hospitalization and/or death was evaluated. For ease of interpretation, only certain operationalizations are shown in this table.

**Table S8**. Discrimination of best-performing Cox model for hospitalization and/or death among young infants with forced inclusion of a variable denoting the presence of at least one World Health Organization danger sign

|  | **Predictor** | **Prevalence, n (%)^a^** | **Adjusted HR (95% CI)** | ***P-*value** | **C-statistic**  **(95% CI)** | **Cross-validated**  **C-statistic (95% CI)** |
| --- | --- | --- | --- | --- | --- | --- |
| **Additional covariates** | Any perinatal or delivery complication | 864 (5.4) | 1.3 (0.73-2.3) | 0.37 | 0.73 (0.69-0.76) | 0.68 (0.58-0.78) |
|  | Umbilical cord care  Antiseptic  Antiseptic and other   substance(s)  Other substance(s) | 9546 (60)  3241 (20)  941 (5.9) | 1.5 (0.88-2.7)  1.6 (0.83-2.9)  1.6 (0.73-3.6) | 0.13  0.17  0.24 |  |  |
|  | Gestational age at birth (weeks), median (25^th^, 75^th^)^b^ | 39.1 (38.3, 40.1) | 1.1 (1.0-1.2) | 0.039 |  |  |
| **History** | Stuffy nose | 1142 (7.1) | 2.8 (1.8-4.2) | <0.001 |  |  |
|  | Yellow discoloration of skin or eyes | 199 (1.2) | 8.7 (5.4-14) | <0.001 |  |  |
|  | Unusual skin rash or anything abnormal on skin | 183 (1.1) | 2.7 (1.3-5.6) | <0.001 |  |  |
| **Physical exam** | Cough | 178 (1.1) | 5.7 (3.2-10) | <0.001 |  |  |
|  | 1 of any of the 8 WHO danger signs | 83 (0.52) | 6.1 (3.4-11) | <0.001 |  |  |

^a^ Denominator is total number of in-person visits (n=16,023).
^b^ Adjusted hazard ratio inverted so that lower gestational age corresponds to higher risk.

**Table S9.** Discrimination of best-performing Cox model for hospitalization and/or death among young infants limited to baseline additional covariates plus symptoms on history only

|  | **Predictor** | **Prevalence, n (%)^a^** | **Adjusted HR (95% CI)** | ***P-*value** | **C-statistic**  **(95% CI)** | **Cross-validated**  **C-statistic (95% CI)** |
| --- | --- | --- | --- | --- | --- | --- |
| **Additional covariates** | Any perinatal or delivery complication | 864 (5.4) | 1.2 (0.69-2.2) | 0.49 | 0.70 (0.67-0.74) | 0.67 (0.57-0.78) |
|  | Umbilical cord care  Antiseptic   Antiseptic   and other   substance(s)  Other   substance(s) | 9546 (60)  3241 (20)  941 (5.9) | 1.6 (0.92-2.8)  1.5 (0.79-2.8)  1.6 (0.71-3.5) | 0.096  0.23  0.26 |  |  |
|  | Gestational age at birth (weeks), median (25^th^, 75^th^)^b^ | 39.1 (38.3, 40.1) | 1.1 (1.0-1.2) | 0.03 |  |  |
| **History** | Stuffy nose | 1142 (7.1) | 2.7 (1.7-4.2) | <0.001 |  |  |
|  | Cough | 691 (4.3) | 3.4 (2.1-5.6) | <0.001 |  |  |
|  | Yellow discoloration of skin or eyes | 199 (1.2) | 11 (6.7-17) | <0.001 |  |  |
|  | Unusual skin rash or anything abnormal on skin | 183 (1.1) | 2.4 (1.2-5.1) | 0.017 |  |  |

^a^ Denominator is total number of in-person visits (n=16,023).
^b^ Adjusted hazard ratio inverted so that lower gestational age corresponds to higher risk.

**Table S10.** Discrimination of best-performing Cox model for hospitalization and/or death among young infants limited to clinical features (with additional covariates removed)

|  | **Predictor** | **Prevalence, n (%)^a^** | **Adjusted HR (95% CI)** | ***P-*value** | **C-statistic**  **(95% CI)** | **Cross-validated**  **C-statistic (95% CI)** |
| --- | --- | --- | --- | --- | --- | --- |
| **History** | Stuffy nose | 1142 (7.1) | 3.2 (2.1-4.8) | <0.001 | 0.67 (0.64-0.71) | 0.66 (0.58-0.75) |
|  | Yellow discoloration of skin or eyes | 199 (1.2) | 11 (7.0-17) | <0.001 |  |  |
|  | Unusual skin rash or anything abnormal on skin | 183 (1.1) | 2.5 (1.2-5.0) | 0.015 |  |  |
| **Physical exam** | Cough | 178 (1.1) | 7.5 (4.3-13) | <0.001 |  |  |

^a^ Denominator is total number of in-person visits (n=16,023).

**Table S11.** Discrimination of Cox model for hospitalization and/or death among young infants (using prevalence of predictors ≥1%) with removal of jaundice hospitalizations

|  | **Predictor** | **Prevalence, n (%)^a^** | **Adjusted HR (95% CI)** | ***P*-value** | **C-statistic**  **(95% CI)** | **Cross-validated**  **C-statistic (95% CI)** |
| --- | --- | --- | --- | --- | --- | --- |
| **Additional covariates** | Umbilical cord care   Antiseptic  Antiseptic and other   substance(s)  Other substance(s) | 9718 (60)  3275 (20)  966 (5.9) | 1.7 (0.93-3.3)  1.7 (0.83-3.4)  1.5 (0.6-3.9) | 0.083  0.15  0.35 | 0.68 (0.67-0.74) | 0.64 (0.53-0.75) |
| **History** | Stuffy nose | 1164 (7.2) | 3.2 (2.1-5.0) | <0.001 |  |  |
|  | Cumulative sum of visits with red, oozing and/or swollen eyes up to the last visit, median (range)^b^ | 0 (0, 8) | 0.43 (0.16-1.1) | 0.092 |  |  |
|  | Yellow discoloration of skin or eyes | 220 (1.4) | 2.3 (0.85-6.4) | 0.11 |  |  |
| **Physical exam** | Cough | 184 (1.1) | 8.2 (4.8-14) | <0.001 |  |  |

^a^ Denominator is total number of in-person visits (n=16,270).
^b^ Value for first visit missing by default due to lagged variable and imputed with 0.

**Table S12**. Discrimination of Cox model for hospitalization and/or death among young infants (using prevalence of predictors ≥0.1% instead of ≥1%)

|  | **Predictor** | **Prevalence, n (%)^a^** | **Adjusted HR (95% CI)** | ***P-*value** | **C-statistic (95% CI)** | **Cross-validated C-statistic**  **(95% CI)** |
| --- | --- | --- | --- | --- | --- | --- |
| **History** | Stuffy nose | 1142 (7.1) | 2.6 (1.6-4.0) | <0.001 | 0.71 (0.67-0.76) | 0.66 (0.57-0.75) |
|  | Yellow discoloration of skin or eyes | 199 (1.2) | 8.8 (5.3-15) | <0.001 |  |  |
|  | Fast or difficult breathing | 32 (0.20) | 11 (3.8-30) | <0.001 |  |  |
|  | Weak, abnormal or absent cry | 9 (0.056) | 28 (6.9-118) | <0.001 |  |  |
| **Physical exam** | Cough | 178 (1.1) | 4.7 (2.5-8.6) | <0.001 |  |  |
|  | Skin rash | 127 (0.79) | 3.7 (1.5-9.0) | 0.0039 |  |  |
|  | Audible wheeze or whistling breath sounds | 89 (0.56) | 2.7 (1.2-6.0) | 0.015 |  |  |
|  | Skin pustules | 43 (0.27) | 2.5 (0.72-8.7) | 0.15 |  |  |
|  | Tachypnea (RR≥60 on 2 repeat measurements) | 19 (0.12) | 0.24 (0.050-1.2) | 0.074 |  |  |
|  | Swollen eyes in the past 7 days | 33 (0.21) | 3.0 (0.82-11) | 0.097 |  |  |
|  | Not sucking effectively | 17 (0.11) | 5.5 (1.8-18) | 0.0036 |  |  |
|  | Severe lower chest wall in-drawing in the past 7 days | 21 (0.13) | 5.6 (1.7-18) | 0.0049 |  |  |
|  | Yellow discoloration of hands and soles of feet | 13 (0.08) | 5.3 (1.9-14) | 0.0011 |  |  |
|  | Fever (one temperature measurement ≥38^o^C or two measurements ≥37.5^o^C) | 11 (0.069) | 22 (6.2-74) | <0.001 |  |  |

^a^ Denominator is total number of in-person visits (n=16,023).

Note: To adhere to the rule of thumb of 10 events per predictor, the following were performed in the stated order: 1) predictors with VIF ≥5 were removed, 2) predictors with p-value >0.001 were removed, 3) ‘Yellow discoloration of skin’ and ‘yellow discoloration of eyes’ jaundice variables were removed because ‘yellow discoloration of hands and soles of feet’ was already included and was the most significant jaundice variable.

**Table S13.** Discrimination of Cox model for hospitalization and/or death among young infants (using prevalence of predictors ≥1%) using only predictors retained after backward selection without manually forcing additional covariates into the model to improve model discrimination

|  | **Predictor** | **Prevalence, n (%)^a^** | **Adjusted HR (95% CI)** | ***P*-value** | **C-statistic (95% CI)** | **Cross-validated C-statistic**  **(95% CI)** |
| --- | --- | --- | --- | --- | --- | --- |
| **Additional covariates** | Gestational age at birth (weeks), median (25^th^, 75^th^)^b^ | 39.1 (38.3, 40.1) | 1.1 (1.0-1.2) | 0.030 | 0.69 (0.66- 0.73) | 0.68 (0.57-0.78) |
| **History** | Stuffy nose | 1142 (7.1) | 2.6 (1.7-4.1) | <0.001 |  |  |
|  | Cough | 691 (4.3) | 1.6 (0.82-3.1) | 0.17 |  |  |
|  | Feels hot to touch or has fever within the past 7 days | 508 (3.2) | 1.7 (0.88-3.2) | 0.12 |  |  |
|  | Runny nose | 227 (1.4) | 2.3 (1.2-4.4) | 0.012 |  |  |
|  | Cumulative sum of visits with red, oozing and/or swollen eyes up to the last visit, median (range)^c^ | 0 (0, 8) | 0.40 (0.15-1.1) | 0.072 |  |  |
|  | Yellow discoloration of skin or eyes | 199 (1.2) | 11 (6.8-17) | <0.001 |  |  |
| **Physical exam** | Skin rash | 127 (0.79) | 5.1 (2.5-11) | <0.001 |  |  |
|  | Axillary temperature (^o^C), median (25^th^, 75^th^)^d^ | 36.3 (36.1, 36.5) | 1.6 (0.96-2.6) | 0.070 |  |  |
|  | Cough | 178 (1.1) | 4.7 (2.3-9.8) | <0.001 |  |  |

^a^ Denominator is total number of in-person visits (n=16,023).
^b^ Adjusted hazard ratio inverted so that lower gestational age corresponds to higher risk.
^c^ Value for first visit missing by default due to lagged variable and imputed with 0.
^d^ Mean of at least two and up to three temperature measurements during the same visit.

**Table S14.** Sensitivity and subgroup analyses for discrimination of best-performing Cox model for hospitalization and/or death among young infants (using prevalence of predictors ≥1%)

| **Analysis** | **Number of events in the analysis** | **C-statistic (95% CI)** | **Cross-validated C-statistic (95% CI)** |
| --- | --- | --- | --- |
| **Best-performing Cox model (using prevalence of predictors ≥1%)** | **174** | **0.71 (0.68-0.75)** | **0.68 (0.58-0.78)** |
| **Sensitivity analyses** | | | |
| Using p-value threshold of <0.05 instead of <0.2 for backward selection | 174 | 0.70 (0.67-0.73) | 0.68 (0.57-0.78) |
| Excluding infants who ever had systemic antibiotic administration during routine home visit assessments | 146 | 0.69 (0.65-0.73) | 0.67 (0.56-0.79) |
| Including hospitalizations and deaths up to 4 weeks after the last CHRW visit | 193 | 0.71 (0.68-0.75) | 0.67 (0.57-0.78) |
| Excluding hospitalization events <48 hours duration | 163 | 0.72 (0.69-0.76) | 0.69 (0.58-0.79) |
| Imputing missing values of time-varying predictors using last observation carried forward^a^ | 176 | 0.70 (0.67-0.75) | 0.68 (0.58-0.78) |
| **Subgroup analyses** | | | |
| 0 to 28 days age group | 118 | 0.70 (0.67-0.76) | 0.65 (0.52-0.78) |
| 28 to 60 days age group | 66 | 0.72 (0.67-0.79) | 0.66 (0.49-0.84) |
| Female | 85 | 0.70 (0.66-0.75) | 0.64 (0.49-0.79) |
| Male | 89 | 0.75 (0.70-0.80) | 0.69 (0.54-0.84) |
| None up to complete primary school | 58 | 0.71 (0.66-0.78) | 0.64 (0.44-0.83) |
| Secondary school incomplete | 50 | 0.73 (0.68-0.81) | 0.69 (0.51-0.87) |
| Secondary school complete or higher | 66 | 0.71 (0.65-0.78) | 0.66 (0.50-0.83) |

^a^ All time-fixed predictors had 0% missingness except for gestational age, which had missingness 1.05%. This missingness was imputed with single imputation using the median gestational age across all infants.

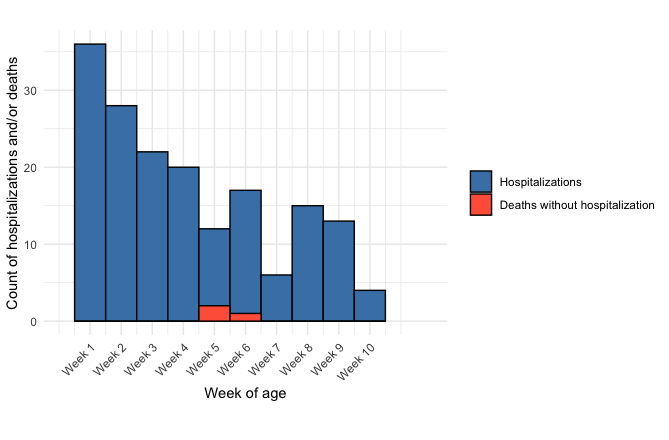

**Figure S1.** Distribution of hospitalizations and deaths in infants who were never hospitalized by week of age

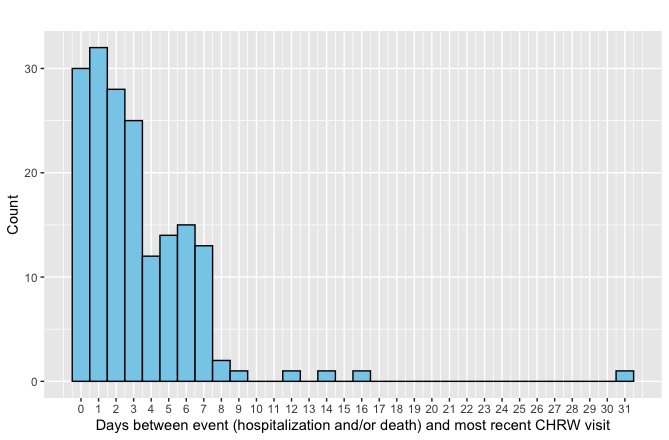

CHRW: Community health research worker.

Overall median (25^th^, 75^th^) days between event and most recent CHRW visit: 2 (1, 5).
Median (25^th^, 75^th^) days between event and most recent CHRW visit for events occurring at 0-30 days of age: 2 (1, 4).

Median (25^th^, 75^th^) days between event and most recent CHRW visit for events occurring at 31-67 days of age: 3 (2, 5).

**Figure S2.** Frequency of durations in days between event (hospitalization and/or death) and most recent community health research worker visit (n=176)

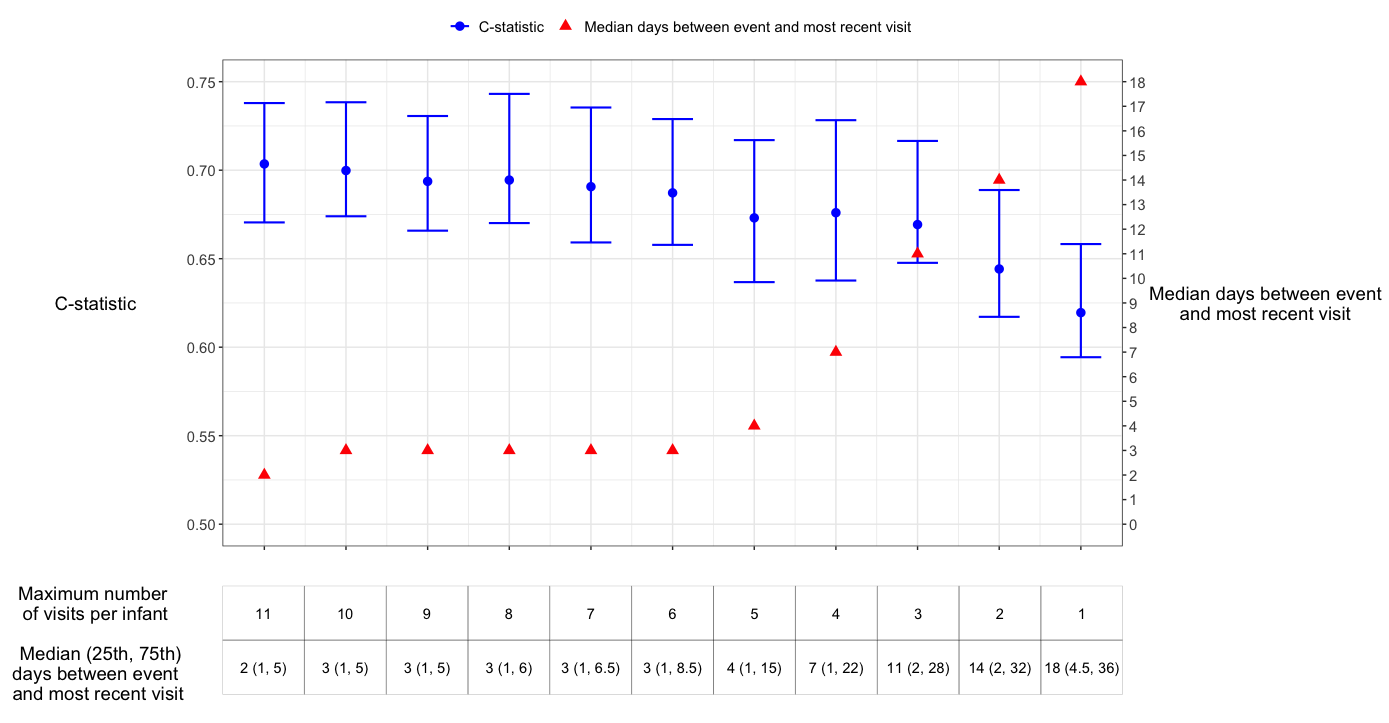

Note: Analyses were restricted to the first *k* scheduled visits per infant, with the number of events held constant across analyses (n=176).

**Figure S3**. C-statistic by maximum number of visits per infant for best-performing Cox model for hospitalization and/or death among young infants (using prevalence of predictors ≥1%)

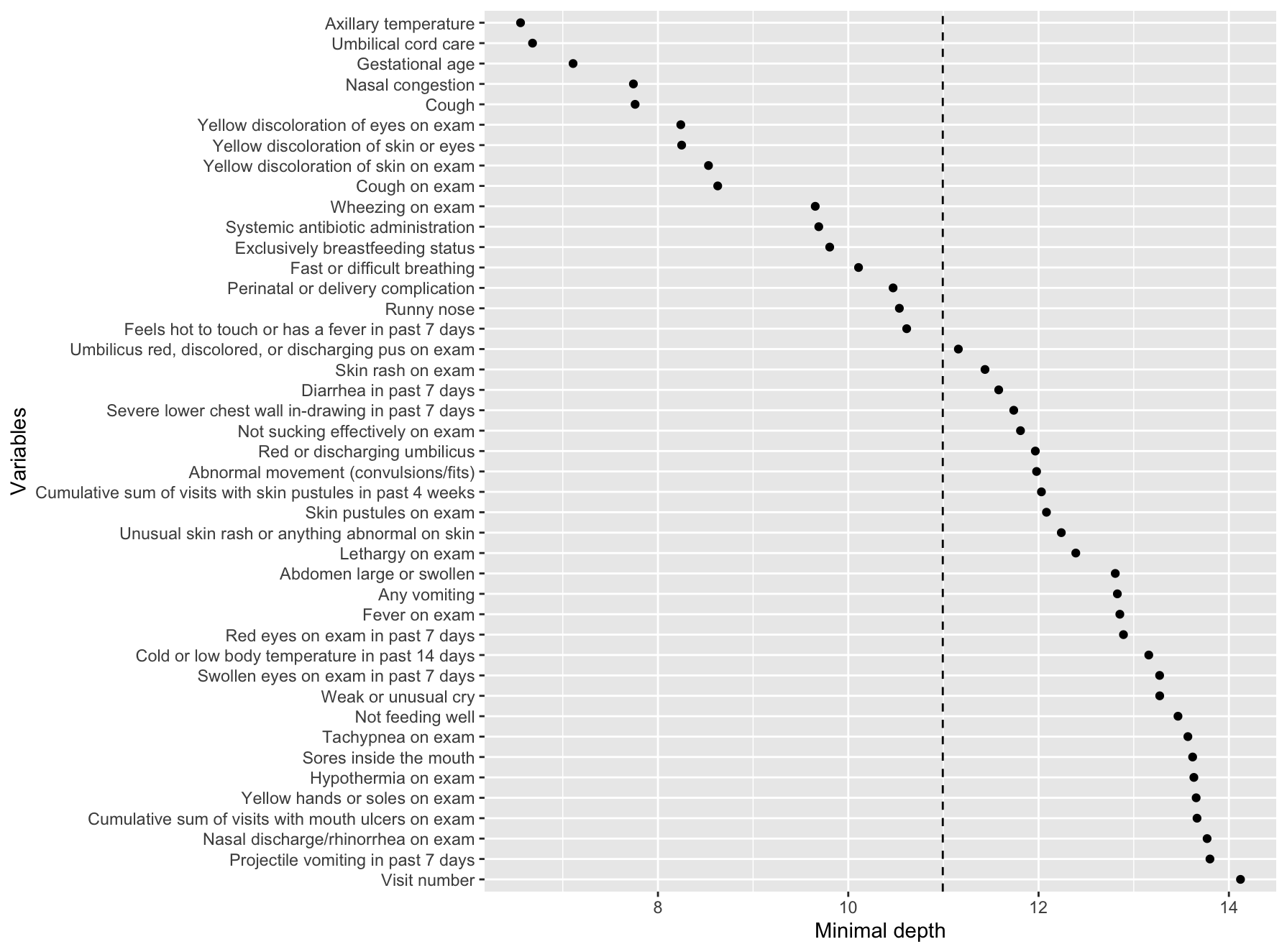

- - - - - - Depth threshold: 10.99

Predictors included in the best-performing time-varying Cox model are highlighted in blue.

WHO danger signs are highlighted in yellow.

**Figure S4.** Variable importance by minimal depth for best-performing Random Forest for Survival, Longitudinal, and Multivariate data (RF-SLAM) model for hospitalization and/or death among young infants (using prevalence of predictors ≥0.1%)

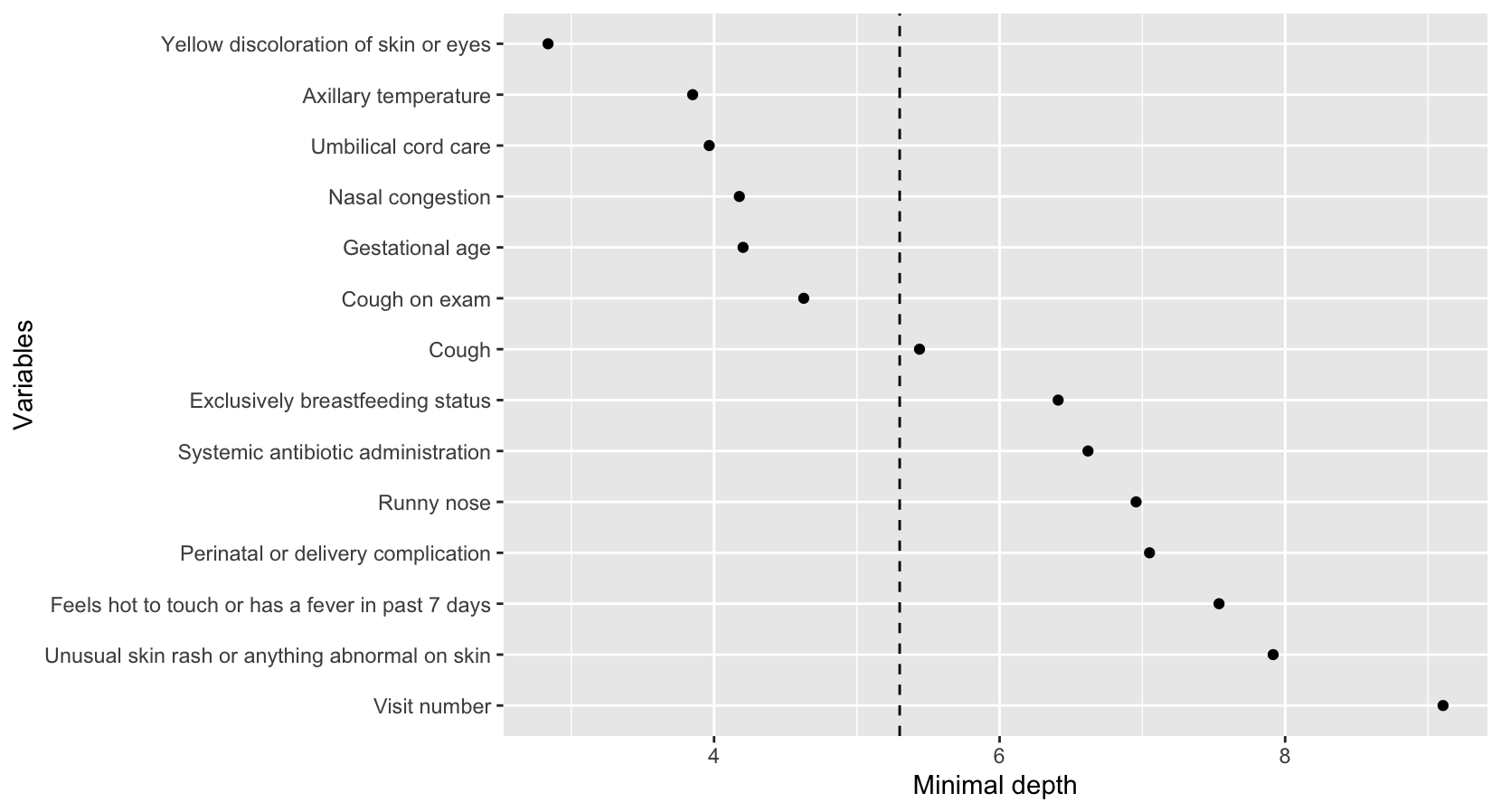

- - - - - - Depth threshold: 5.30

**Figure S5**. Variable importance by minimal depth for Random Forest for Survival, Longitudinal, and Multivariate data (RF-SLAM) model for hospitalization and/or death among young infants (using prevalence of predictors ≥1% instead of ≥0.1%)

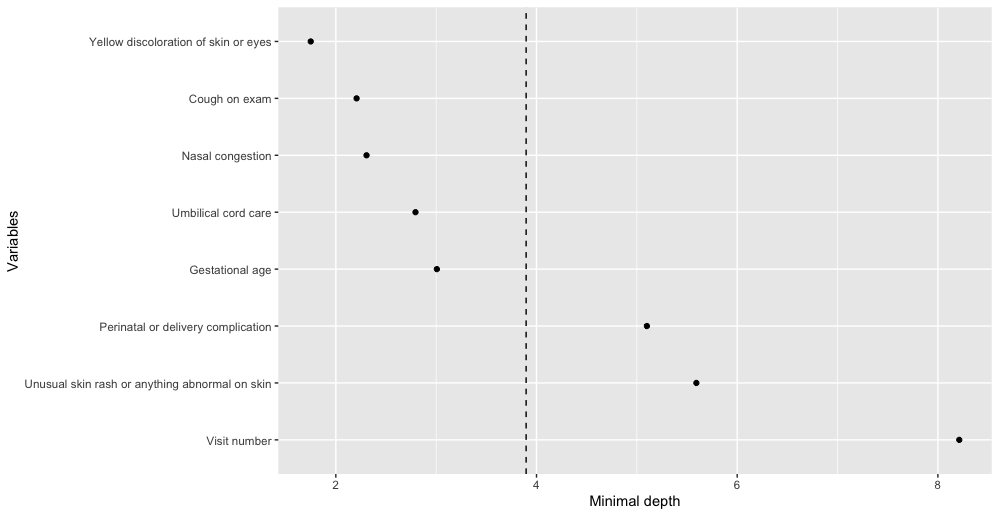

- - - - - - Depth threshold: 3.90

**Figure S6**. Variable importance by minimal depth for Random Forest for Survival, Longitudinal, and Multivariate data (RF-SLAM) model for hospitalization and/or death among young infants using predictors from best-performing time-varying Cox model

**References Cited**

1.	World Health Organization. Acute respiratory infections in children: Case management in small hospitals in developing countries. World Health Organization. <https://apps.who.int/iris/bitstream/handle/10665/61873/WHO_ARI_90.5.pdf>

2.	World Health Organization. WHO recommendations on postnatal care of the mother and newborn. World Health Organization. <https://apps.who.int/iris/bitstream/handle/10665/97603/9789241506649_eng.pdf?sequence=1>
